# Effective Reproduction Number and Dispersion under Contact Tracing and Lockdown on COVID-19 in Karnataka

**DOI:** 10.1101/2020.08.19.20178111

**Authors:** Siva Athreya, Nitya Gadhiwala, Abhiti Mishra

## Abstract

We analyze the data provided in the Novel Coronavirus (COVID-19) media bulletins of the Government of Karnataka. We classify the patients of COVID-19 into clusters and study the Reproduction number and Dispersion for eight specific clusters. We find that it is uniformly less than one, indicating the benefits of contact tracing, lockdown and quarantine measures. However, the Dispersion is low indicating individual variation in secondary infections and the occurrence of Super-spreading events. Finally, we analyze the surge in infections after 27th June and find it unlikely that it was caused solely by the large Migration in May and June 2020.

## 1 Introduction

For COVID-19, in the absence of a vaccine, key measures to contain infection spread have been lockdowns, contact tracing, quarantine, testing along with wide publicity of social distancing norms, hygiene guidelines, awareness of the symptoms of the disease and treatment. There are many efforts to understand control measures such as lockdowns, contact tracing and quarantine with respect to COVID-19 spread using stochastic models. In [1], using a stochastic transmission model it has been concluded that highly effective contact tracing and case isolation is enough to control a new outbreak of COVID-19 within 3 months and that the probability of control decreases with long delays from symptom onset to isolation, fewer cases ascertained by contact tracing, and increasing transmission before symptoms. In [2], the authors seem to suggest that COVID-19 spreads too fast to be contained by manual contact tracing, but could be controlled by a contact-tracing app which is faster, more efficient, and on a larger scale. They claim that by targeting recommendations to only those at risk, epidemics could be contained without resorting to lockdowns.

Contact tracing and other controlled measures were also used by countries during the Severe Acute Respiratory Syndrome (SARS) epidemic. In [3], the authors use detailed epidemiological data from Singapore and epidemic curves from other settings, to estimate reproductive number for SARS in the absence of interventions and in the presence of control efforts. They conclude that a single infectious case of SARS infects about three secondary cases in a population that has not yet instituted control measures. In [4], the authors study the first 10 weeks of the SARS epidemic in Hong Kong. The epidemic was characterized by two large clusters-initiated by two separate “super-spread” events (SSEs)—and by ongoing community transmission. Using a stochastic model, they compute basic reproduction number and transmission rates and conclude that the result of reductions in population contact rates, improved hospital infection control and rapid hospital attendance by symptomatic individuals resulted in fall of transmission rate and decline of the epidemic.

In [5], they argue that using only the basic reproduction number can obscure the individual variation in infectiousness. Their motivation being ‘super-spreading events’- in which certain individuals had infected unusually large numbers of secondary cases (5–10 in the SARS epidemic). They studied contact tracing data from eight directly transmitted diseases, and showed that the distribution of individual infectiousness around the basic reproduction number is skewed. Using various models they then proceed to compare effect of individual-specific control measures versus population-wide measures. They conclude that super-spreading events are a normal feature of disease spread and give a formal definition of the same.

In [6], the authors study the epidemiology and transmission of COVID-19 in Tamil Nadu and Andhra Pradesh using testing and contact-tracing data. They conclude that 92.1% of cases and 59.7% of deaths occur among individuals less than 65 years old. They use the timeline and the mortality data along with the EpiEstim package to estimate the reproduction number. They also point to the fact that Superspreading plays a prominent role in transmission, with 5.4% of cases accounting for 80% of infected contacts. They also estimate the per-contact risk of infection, case-fatality ratio and median time-to-death.

Since 9^th^ March 2020, the Government of Karnataka has been providing detailed media bulletins (click here for bulletins till April 26^th^ 2020 and here from April 27^th^, 2020) containing specific guidelines on the virus and information on each patient who was tested positive in the state. The bulletins on the tested positive patients contain information regarding how each one of them contracted the virus (either due to travel history or by being a contact of someone who has already tested positive for COVID-19) or what led to them being tested (either as a Severe Acute Respiratory Infection patient or someone with Influenza like symptoms).

In this article we study the trace history provided in the media bulletins and try to understand the spread of the disease in the period from 9^th^ March till 21st July 2020 in the state of Karnataka. From the trace history classify the patients who tested positive into several clusters. We analyse each cluster and the spread of disease within them. We also comment on the reasons for the possible spurt in cases from 27th June, 2020 onwards.

### 1a Remark

*A word of caution before we proceed. Our entire work is based on data provided in the Media Bulletins, [7]. These are dependent on the contact tracing procedures and testing policy followed by the government, many details of which are unknown to us. From Figure 1, the fraction of positive tests is around 6.77% at this time and it is the highest fraction recorded. On 15^th^ May, 2020 it reached an all-time low of 0.7%. The number of total tests conducted up to 21^st^ July is 1049982 which include RAT, RT-PCR and other testing techniques. The details as to the amount of tests done using each technique was not mentioned before 17^th^ July. These provide a comprehensive count of testing numbers in the state but not cluster wise testing data.*

**Figure 1:**
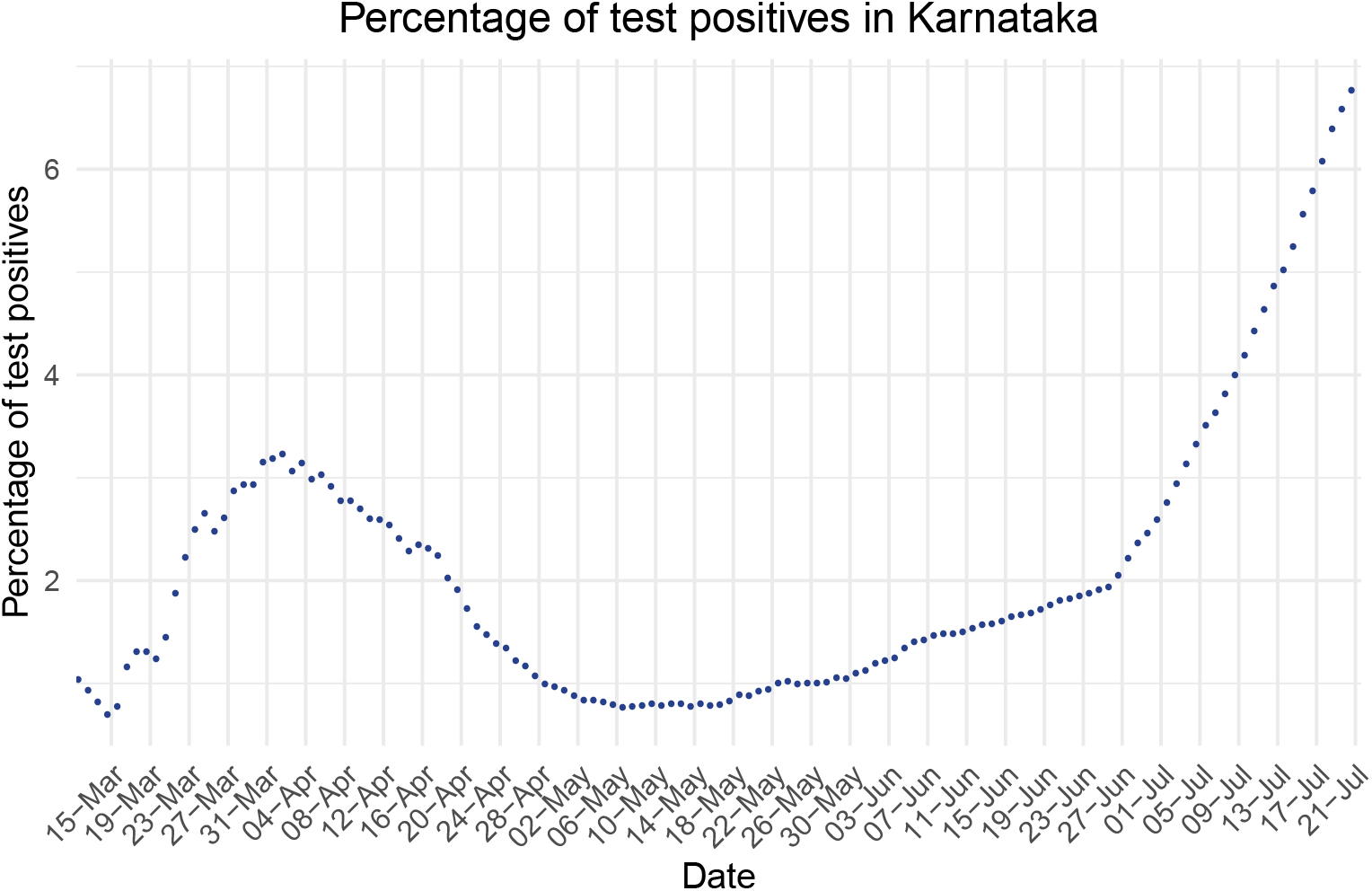
The above graph represents the percentage of test positives in Karnataka. For each day, we plot the percentage of the total positives up to that day over the total number of tests conducted up to that day.

*Needless to say, the number of infected individuals in the population differs from the number of positive test results. So equating the number of those tested positive to the number of infected individuals may be an error, because every individual in the population has not been tested. Thus for any inference or conclusion on true infection growth we must take into consideration the different policy/rates of testing, population density, contact tracing, quarantine measures, and biological aspects of this epidemic.*

The rest of the paper is organised as follows: In Section 2 we describe our classification of COVID-19 patients in Karnataka into clusters. For eight specific clusters, we fit a Negative Binomial model, (i.e. a mixture of Gamma and Poisson), estimating the basic reproduction number and the dispersion parameter (along with its 95% confidence intervals). In 124 Section 3, we present our conclusions detailing variations in basic reproduction number of the clusters with respect to time, age, generation and districts. We also analyze the migration cases from May and June and conclude that it is unlikely that they are the sole cause for the surge in cases in the month of July. In the Appendix: we begin in Section A by describing the Negative Binomial model used for each cluster’s secondary infection distribution; in Section B we discuss the Maximum Likelihood Estimator for dispersion and reproduction number; in Section C the *χ*^2^ goodness of fit test; and in Section D we describe the methodology to estimate confidence intervals for dispersion.

## 2 Clusters, Reproduction number, and Dispersion

In Karnataka, from the very beginning, quarantine measures and contact tracing were put in place for all tested positive patients. To contain the spread of COVID-19 infections in India, the Union Government started a strict lockdown on 25^th^ March and relaxed them over 5 phases as follows: Lockdown Phase 1 (25^th^ March–14^th^ April) and Lockdown Phase 2 (15^th^ April– 3^rd^ May) were the strictest in terms of mobility; Lockdown Phase 3 (4^th^ May – 17^th^ May) and Lockdown Phase 4 (18^th^ May – 31^st^ May) included relaxations of travel between states; and Unlock 1.0 (1^st^–30^th^ June), Unlock 2.0 (1^st^–31^st^ July) have had considerable relaxations.

The daily media bulletins provided by the Karnataka State Ministry of Health and Family Welfare, apart from issuing regular guidelines for the public, contain detailed information of screening of air/sea passengers, people under observation, number of positive tests on each day and details of each infected patient [including: age, sex, district, source of infection, etc.] We first classify the cases into clusters based on the source of infection, for example “From Europe” or “Pharmaceutical Company Nanjangud”. Then in each cluster we place all the patients who contracted the virus independently from the place of origin, and then recursively add the patients who they passed the infection to.

### 2.1 Clusters

Before Phase 1 (25^th^ March – 14^th^ April) of the lockdown began, almost all the COVID-19 cases that were confirmed in Karnataka were either individuals who had some form of international travel history (from Middle East, USA, South America, United Kingdom and the rest of Europe) or those who were contacts of such individuals. These initial infections were very well contained as many of the infected individuals were confirmed and isolated quite early. Most of these clusters showed very few generations and very few infections were caused by the individuals in these clusters. Phase 1 and Phase 2 (15^th^ April – 3^rd^ May) of the lockdown in Karnataka saw heavy restrictions on travel and nearly all services and factories were suspended.

During Phase 1 and Phase 2 of the lockdown, a Pharmaceutical company in Nanjangud, Mysore, saw a sudden increase in the COVID-19 cases. Although the exact reason for the infection to have reached the company is unknown, the first patient to be infected (35 year old male, was confirmed to be infected on 26^th^ March) came in contact with health care workers treating COVID-19 patients. Another cluster that began during this period was the “TJ Congregation”, which contained those who attended the TJ Congregation from 13^th^ to 18^th^ March in Delhi. The first patient in this cluster was confirmed as a COVID-19 case on 2^th^ April. Both these clusters were very well contained and the last patients to be attributed to these clusters tested positive on 29^th^ April and 21^st^ May respectively. No more patients were attributed to these clusters since then. Phase 3 and 4 of the lockdown loosened restrictions on Domestic Travel and many infected individuals had some domestic travel history. The state saw a large influx of infected individuals from states like Maharashtra, Gujarat, Rajasthan and the Southern States (Tamil Nadu, Telengana and Andhra Pradesh). There were also patients whose source of infection was listed as inter-district travel in Karnataka, travel to foreign countries or other states, healthcare workers and policemen on COVID-19 duty and their contacts. The cases due to these reasons were too few to form separate clusters. We placed all these patients in a cluster called “Others”.

Testing strategy in India is governed by ICMR guidelines. The guidelines on 20th March mandated that all Severe Acute Respiratory Illness patients (i.e. patients with fever AND cough and/or shortness of breath) should be tested for COVID-19, while the guidelines on 4^th^ April mandated the same for all symptomatic patients with Influenza like Illness (fever, cough, sore throat, runny nose). Thus two other clusters that began during Phase 1 and Phase 2 of the lockdown were the Severe Acute Respiratory Infection (“SARI”) (first infection 7^th^ April) and Influenza Like Illness (“ILI”) (first infection 15^th^ April) clusters. These clusters contain those patients who have a history of SARI(and ILI), and those who can be traced back as contacts of such patients. It should be noted that only the first generation of the patients in this cluster are those with a history of SARI (and ILI), but the subsequent contacts of these patients need not be. In the media bulletins, patients whose contact tracing was incomplete were mentioned as ‘Contact Under Tracing’. We have assumed that these patients did not fall under SARI or ILI and placed them in a cluster called “Unknown”, along with their contacts who tested positive. An initiative taken by the government was to create Containment Zones in certain regions. The guidelines for these zones were clearly specified. The first case in contact with a containment zone was reported on 24^th^ April. Since then a large fraction of the increase in this cluset occurred during Phase 3 (4^th^ May – 17^th^ May) and Phase 4 (18^th^ May – 31^st^ May) of the lockdown. For all these clusters, there was no information provided on the source of infection for the ‘parents’.

Please refer to Table 1 for the entire list of clusters.

**Table 1:** From the reasons given for the source of infection, we have classified the patients who tested positive for COVID-19 in Karnataka into distinct clusters.

| Classification of Patients into Clusters |  |
| --- | --- |
| • From Middle East<br>• From USA<br>• From United Kingdom<br>• From Rest of Europe<br>• From South America<br>• From Maharashtra<br>• From Rajasthan<br>• From Southern States<br>• From Gujarat | • Influenza like illness (ILI)<br>• Severe Acute Respiratory Infections(SARI)<br>• Unknown<br>• Pharmaceutical Company-Nanjangud<br>• T.J. Congregation in Delhi<br>• Containment Zones<br>• Others |

In the interactive graph, on our accompanying website (see [8], we plot the trace history as a tree like graph. In the graph of each cluster, the first generation nodes [at depth one] are the patients who got the infection directly from the place of origin of the infection. These patients will be called ‘parents’ of the cluster. The ‘children’ are the people who contracted the disease from the people labelled as ‘parents’, that is, they are at a depth of two in the trace history chart. Similarly, ‘grandchildren’ and ‘great grandchildren’ have depth three and four respectively. The ‘parents’, ‘children’, ‘grandchildren’ and so on have also been referred to as ‘Generation 1,’ ‘Generation 2,’ ‘Generation 3’ and so on. A part of the graph can be seen inFigure 2.

**Figure 2:**
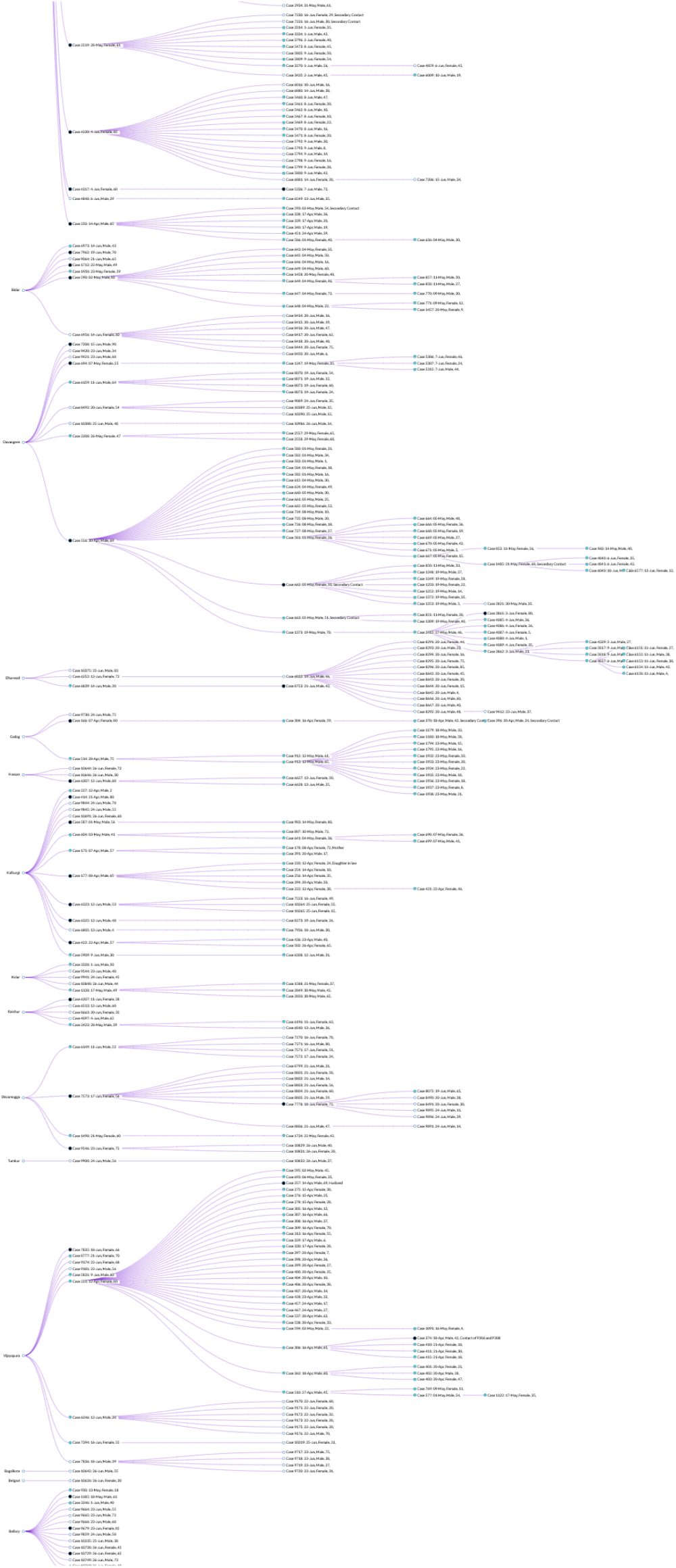
Karnataka Trace History

On a related observation, suppose we consider the entire tested positive population in the state and examine the distribution of the number of infections designated as contacts of earlier infections (see Figure 3). In Figure 3, a large peak is seen at 0 infections caused. It can be seen that only 9 individuals in the population of 4895 have passed the infection on to more than 20 people. This could be the result of a super-spreader phenomenon or perhaps an effect of how the contact tracing and testing is performed. Assigning them as definitely arising from one particular individual will need a more careful understanding of the latter. One can further note that due to effective quarantine measures there are 4265 infected individuals who have not passed the infection on to anyone else.

**Figure 3:**
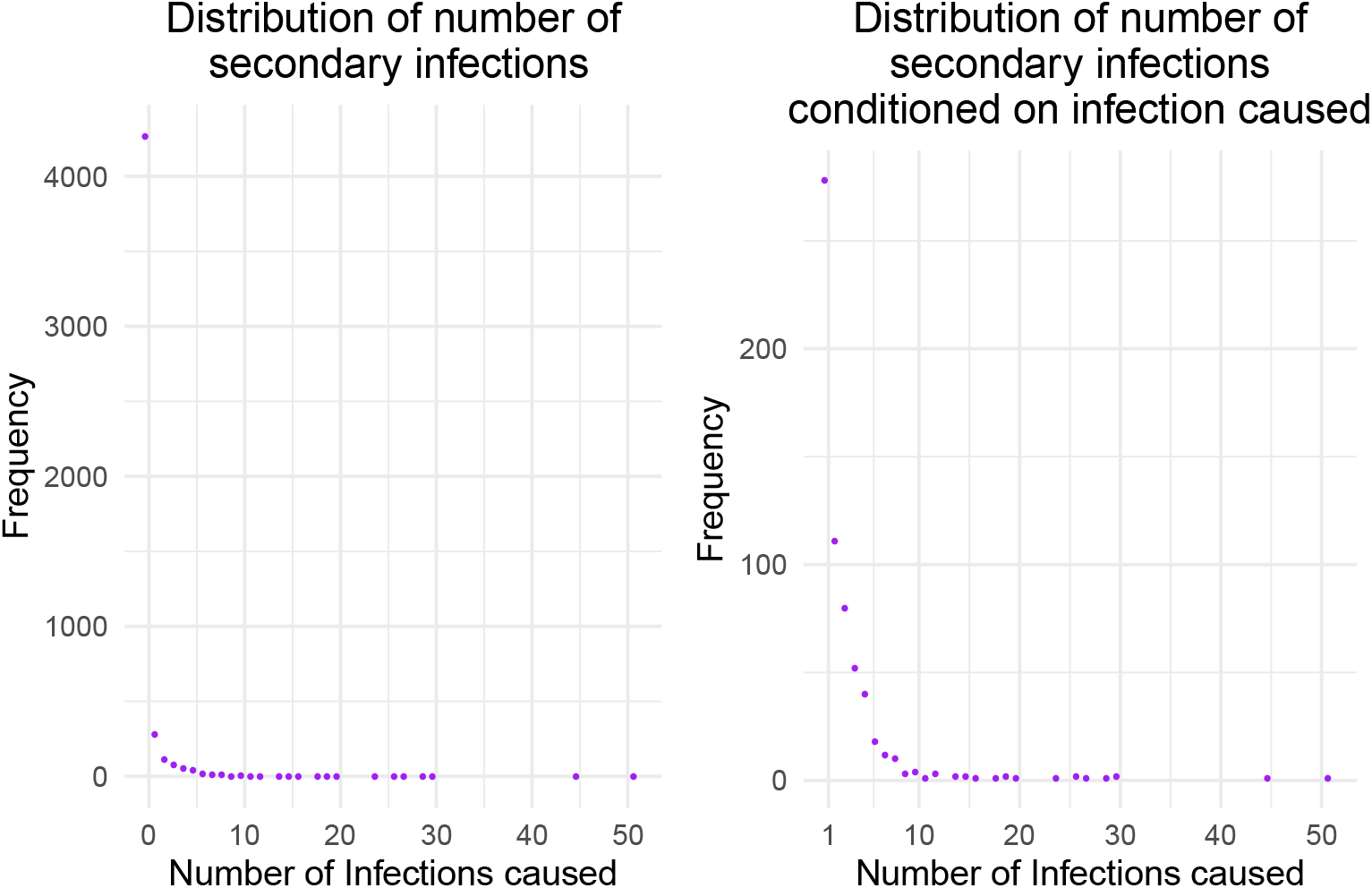
The above scatter plot considers the COVID-19 patients in Karnataka and represents the distribution of the number of infections caused by each patient. The patients belonging to “The 8 clusters” in study are considered here. The plot on the left shows the frequency distribution of the number of infections assigned to each infected individual as their contact. The plots above have the number of infections caused on the *x*-axis and the number of patients that have caused *x* many infections on the *y*-axis. The graph on the right is the same as the one on the left without the point at 0. It is the distribution of number of infections caused conditioned on at least one infection caused.

Please note that the trace history is a measure of how contact tracing is being done and how infected individuals are being identified for testing. Many of the contacts tested are contacts of multiple COVID-19 patients and assigning them to any one contact may not be indicative of the way the infection was passed on. It is also important to note that the parent to child relationship in trace history is indicative of the testing policy and contact tracing that was followed and need not be a definitive indicator of the genealogy of the infection spread.

### 2.2 Reproduction number and Dispersion

In epidemiology, the “basic reproduction number” of an infection, denoted by *R*_0_, can be thought of as the expected number of cases to have contracted the infection directly from one case. Thus on an average, each infected person passes on the infection to *R*_0_ many healthy individuals. As mentioned earlier, in Karnataka during the period 9^th^ March – 26^th^ June we have observed the COVID-19 infection spread in a controlled environment. So whenever we calculate basic reproduction numbers we are actually calculating the short term effective reproduction number of the disease during this period. To be cognizant of this we shall use the notation *R*_eff_ to denote the basic reproduction number for a cluster instead of the usual notation *R*_0_.

We will examine Reproduction number and dispersion for “The 8 clusters” in this section, namely

- From Southern States
- Influenza like illness
- Severe Acute Respiratory Infections
- Containment Zones
- Unknown
- Others
- TJ Congregation in Delhi
- Pharmaceutical Company Nanjangud

These began before 3^rd^ May 2020 and have more than 50 individuals in total. There are ten clusters that satisfy this criteria from Table 1. We have omitted two clusters from analysis which satisfy this criteria, namely: “From Maharashtra” and “From Middle East”. We shall explain the reasons for the same in next section.

In Figure 4 we present a summary distribution of parents, children, grandchildren, and great grandchildren in each of “The 8 clusters”. We will now focus on the distribution of children for each of the clusters. For each individual *i* in the cluster we will denote the number of children (or the number of tested positive cases) assigned to patient *i* by *y*_i_. This means that there were *y*_i_ many positive infections who the media bulletins listed as ‘Contact of Patient-*i*’. The mean of *y*_i_ is the basic reproduction number *R*eff. In Table 2 we present a comparison of the summary distributions parameters (Maximum, Zeroes, Size, etc.) across clusters. From Table 2 we see that the variance does not match the mean. Further, as noted in Figure 5, heterogeneity in the infectiousness of each individual implies that *R*eff by itself is not a good measure of the infection spread. To account for the large variance, we now consider the standard method of mixture of Poisson distributions to model the data set. For each cluster, using the Negative Binomial with mean *R*_eff_ and dispersion *k* (see [5] and Section A for details) as the offspring distribution, we will use the Maximum Likelihood method for estimating *R*_eff_ and *k* (see Section B for details).

**Figure 4:**
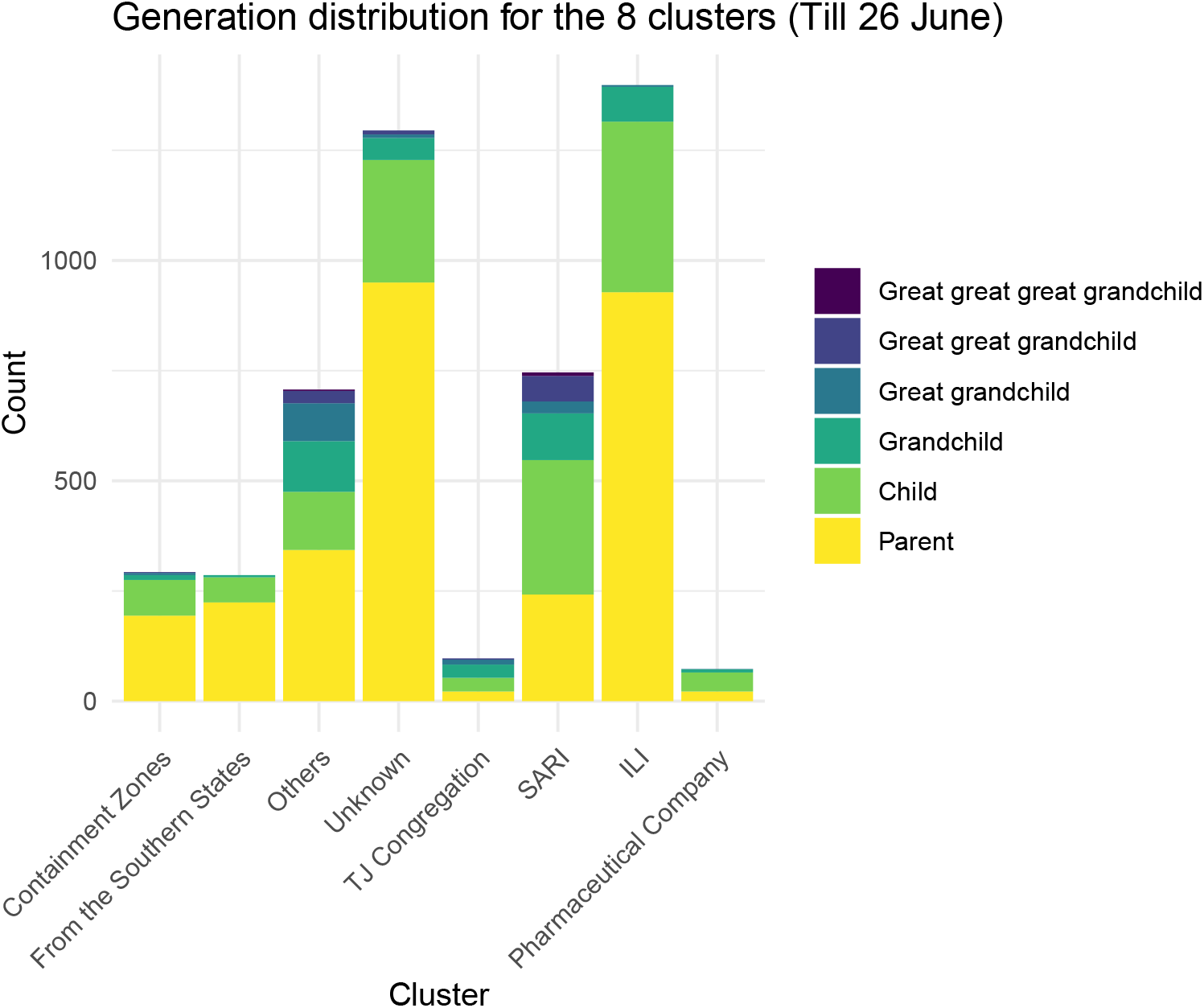
The above plot is a stacked histogram displaying the distribution of generations for“The 8 clusters” we consider till 26^th^ June. The histogram represents the number of infections that belong to each of these clusters and each bar has been further filled with different colors to denote the number of primary infections, secondary infections and so on.

**Table 2:** The above table contains the following information about the 8 clusters that we have considered- the ‘Size’ column represents the number of infected individuals that belong to the cluster. The ‘Zeros’ column denotes the number of patients who haven’t been assigned any secondary infection. The ‘Maximum’ column represents the maximum of the number of secondary infections assigned to any individual. The ‘*R*_eff_’ and ‘Variance’ column represents the mean and variance (respectively) of the secondary infections assigned to individuals.

| Name of Cluster | Size | Zeros | Maximum | $R_{eff}$ | Variance |
| --- | --- | --- | --- | --- | --- |
| Containment Zones | 293 | 254 | 7 | 0.3447 | 1.199 |
| From the Southern States | 286 | 257 | 7 | 0.2168 | 0.6897 |
| ILI | 1398 | 1243 | 30 | 0.3369 | 2.355 |
| Others | 707 | 593 | 51 | 0.5191 | 6.502 |
| Pharmaceutical Company | 73 | 53 | 24 | 0.726 | 8.757 |
| SARI | 746 | 622 | 45 | 0.6743 | 8.521 |
| TJ Congregation | 97 | 70 | 15 | 0.7732 | 3.823 |
| Unknown | 1295 | 1173 | 27 | 0.2625 | 1.637 |

**Figure 5:**
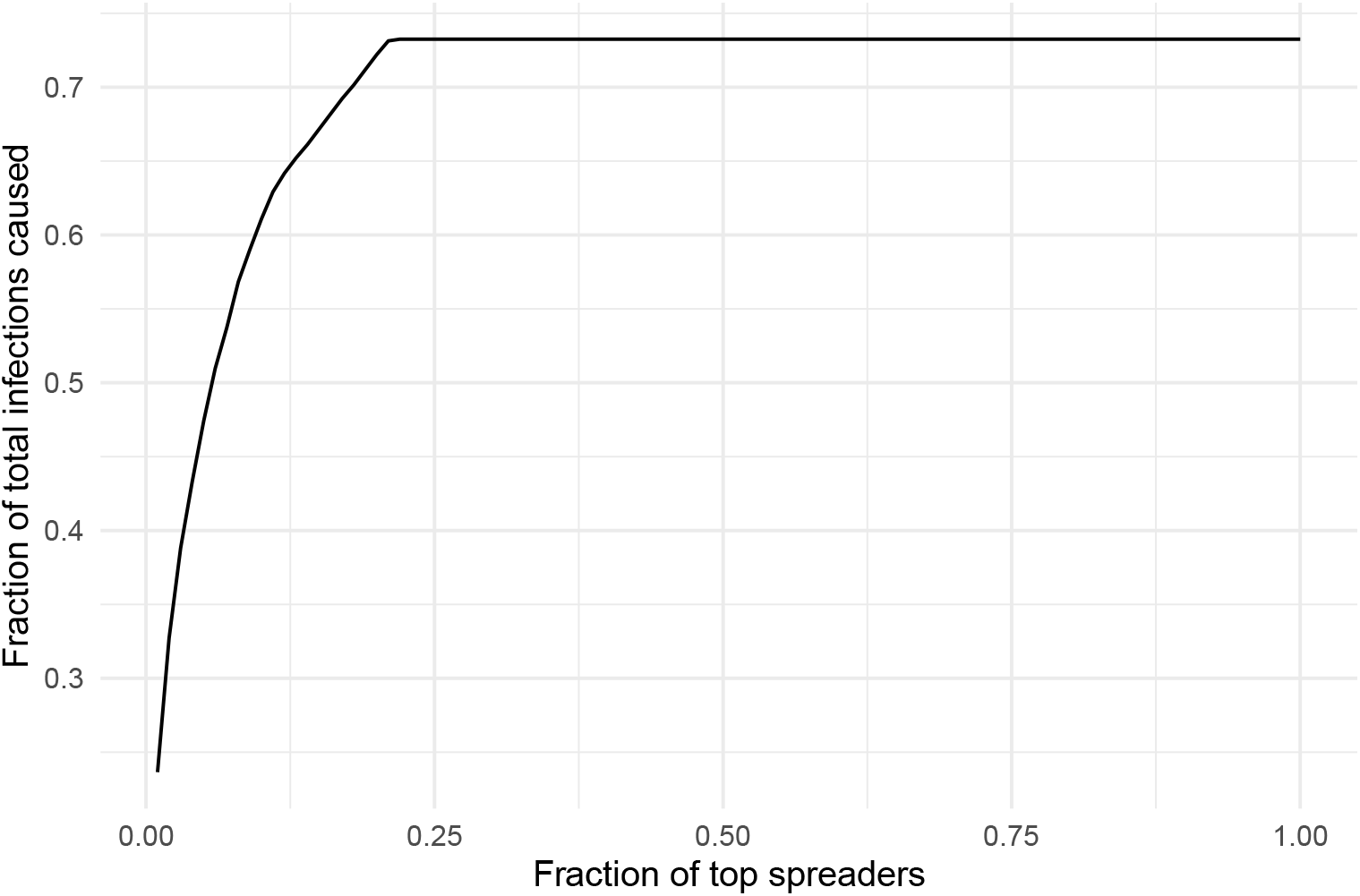
The plot considers those individuals who were infected before 3rd May along with all those cases that can be traced back as contacts of them. The infected individuals have been ranked in terms of the number of secondary infections caused by them. Then the top *x* fraction of them are considered. In the above graph, *x* has been plotted on the x-axis and the fraction of the total infections infected by them on the *y*-axis.

We begin by explaining in detail the analysis for the “Containtment Zone” cluster and then present a summary for the remaining clusters. The size of the cluster is 293, the maximum number of secondary infections assigned to an infected person is 7, there are 254 infected individuals in this cluster who have been assigned 0 secondary infections. The Maximum likelihood estimates for *R*_eff_ and *k* are given by *R*_eff_ = 0.3447 and *k* = 0.09345. The 95% confidence interval for *k* is (0.06736, 0.15250) and the *p*-value from the *χ*^2^-goodness of fit test is given by 0.5966. The methods of calculating the numbers above can be found in Sections B, C and D of the Appendix. As the confidence interval is fairly small and the *p*-value is not less than 0.05, the Negative Binomial model can be selected for the data set.

## 3 Conclusions

From the analysis of “The 8 clusters” we can see the definitive effect of contact tracing, quarantine measures and lockdown, as the mean reproduction numbers were uniformly less than one. Despite best effort of manual contact tracing “SARI” and “ILI” clusters continued to add new ’parents’ indicating presence of viral load in the population. We shall also conclude below that cases due to migration in Phase 3, Phase 4 and Unlockdown 1.0 were unlikely to be the cause of the surge in infections in July, 2020. We shall justify these summary conclusions in this section.

One thumb rule for disease spread, including COVID-19 anecdotally, is the 20/80 rule. The rule states that 80% of the secondary infections arise from 20% of the primary infections. As seen in Figure 5 if we look at patients in Karnataka who tested positive on or before 3rd May and their descendants then the rule holds true. Figure 5 attempts to demonstrate the heterogeneity of the infectiousness of the individuals infected with COVID-19 in Karnataka. It can be observed that for Karnataka, almost 20% of the individuals with the highest infectiousness are responsible for 70% of the total infections. In the case of a perfectly homogeneous population of infected individuals (i.e., every infected individual infects equal number of healthy individuals), would represent the *y* = *x* line. The large deviation from the *y* = *x* straight line represents the heterogeneity in the infected individual population.

### “The 8 clusters”

- **Heterogeneity and Variation**In Table 3, we have computed the Maximum Likelibood estimators for *R*_eff_ and *k* for “The 8 clusters” and also performed the *χ*^2^-goodness of fit test (see Section C for details regarding the goodness of fit). In Figure 6 we have plotted the histogram from the derived Negative Binomial probabilities for each of “The 8 clusters” along with the observed relative frequencies of the number of infections caused. We have marked the 95^th^ and 99^th^ percentile for these distributions in the plot. In Table 4 and Figure 7 we provide the confidence intervals for dispersion parameter with respect to “The 8 clusters”. The “TJ Congregation” and the “Pharmaceutical Company Nanjangud” clusters both have higher *R*_eff_ among all clusters. The *p*-values in the last column of Table 3 are not small for all clusters except for the cluster “Pharmaceutical Company Nanjangud”. This cluster has a very high variation, a maximum data point at 24 (i.e. one person who has been assigned to 24 secondary infections) and also a significant proportion at 1 secondary infection caused. One can also see that the confidence interval for the dispersion for “Pharmaceutical Company Nanjangud” cluster is quiet large as well, as seen in Figure 7 and that the histograms differ with the Negative Binomial model in Figure 6 as noted. For each cluster in “The 8 clusters”, we found that basic reproduction number is less than 1 but the variance is larger than the mean (see Table 2). However, the distribution of secondary infections across all clusters is very skewed, with a significant mass at 0 due to the control measures taken. From the Negative Binomial model,(as proposed in [5] to account for this variation), we note that for most clusters their dispersion is low and is contained in a small confidence interval. Thus, though the clusters will most likely die out under the controlled environment, there is a reasonable chance of super-spreading events occuring.
- **Super-spreading events**
Note that in Table 2, the largest number of secondary infections assigned to an individual is quite high for some clusters. This might be indicative of the super-spreading phenomenon. A general protocol for defining a super-spreading event is given in [5]. It is as follows: (1) estimate the effective reproductive number, *R*_eff_, for the disease and population in question; (2) construct a Poisson distribution with mean *R*_eff_, representing the expected range of *Z* (without individual variation); (3) define a Super-spreading event as any infected individual who infects more than *Z*_n_ others, where *Z*_n_ is the *n*^th^ percentile of the Poisson(*R*_eff_) distribution. If *R*_eff_ and *k* have been estimated then one can use the definition and the Negative Binomial model to understand the probability with which such events will occur. If we were to consider a 99th percentile event with the above *R*_eff_ = 0.3447, then an event causing more than 2 secondary infections would be considered a super-spreading event. In the “Containtment Zone” cluster, there is a person who has been assigned 7 secondary infections, this would be considered a super-spreading event. Under the Negative Binomial model the probability of observing 7 secondary infections is 0.0027. This may indicate one of two possibilities, either a very, very rare event has occurred or it is just an effect of the testing and contact tracing method that was followed. The relative frequency of super-spreading events within “The 8 clusters” can be calculated using Table 5 and Figure 6. The above indicates that the infection can be stemmed quicker by containing these super-spreading events by using effective contact tracing.
- **Variation over time**
If we consider 7^th^ April and Descendants till 21^st^ April, then there were 290 patients who tested positive and 219 out of them did not pass the infection to anyone else. There was one person who had been assigned 24 secondary infections and the mean number of secondary infections was at 0.6793 with a variance of 4.482. In contrast, if we consider the period 7^th^ April to 3rd May and Descendants till 17^th^ May, then there were 615 patients who tested positive and 491 out of them did not pass the infection to anyone else. There was one person who had been assigned 45 secondary infections and the mean number of secondary infections was at 0.7512 with a variance of 9.946. The basic reproduction number is by no means a unique number for a disease or for that matter within a cluster. It greatly varies: with time from begining to end; within a region due to it’s population density; and with interventions put in place to curb the spread of the infection. In Figure 8 we compute the reproduction number for each of “The 8 clusters” studied in Section 2 on a weekly basis. We note that there is a signficant variation over time. The “Pharmaceutical Company” cluster seems to have a reproduction number of 4 during the first week and then tappers off to 0 in five weeks. The “TJ Congregation” cluster also has a reproduction number that has variation over time but eventually due to tracing and testing tappers off to 0. “SARI” and “ILI” clusters have fluctuations throughout the period, due to new parents being added to the cluster. Even a meticulously planned manual contact tracing effort has in-built time constraints. This time delay, when the number of cases rise, may result in super-spreading events mentioned earlier. Also “SARI”/“ILI” clusters indicate some local transmission in the state making complete accounting of secondary infections via manual contact tracing a big challenge.
- **Variation over Generations**Table 6 contains information on “The 8 clusters” with respect to generations within them. The maximum number of infections caused by an individual in the first generation is 30. An individual in the “Influenza like illness” cluster and another in the “Others” cluster have caused 30 secondary infections each. Among the individuals in the second generation the one to have caused 51 infections belongs to the “Others” cluster. It is observed that the mean secondary infections of patients belonging to the Generation-4 (Great Grandchild) is 0.7042 and is significantly higher than the remaining generations. This is because of the small size of the generation (142) and one of the patients being assigned 45 secondary infections. While the highest generation that can be observed is Generation-6 (Great great great grandchild), they haven’t been included in Table 6 as there isn’t a Generation-7 for any cluster resulting in all the individuals in Generation-6 being assigned 0 infections. A heat map representing the mean infections caused, as studied across clusters and generations, is seen in Figure 9. All clusters have been contained within 5 generations till now, this is seen by the fact the mean is 0 for the final generation of the cluster which is at most the fifth. One can see that the “TJ Congregation” and “Pharmaceutical Company Nanjangud” clusters have variation in mean across generation with the mean number of infections decreasing across each generation. The clusters have closed out and have not added any new patients for some time. Generation 3 in the “SARI” cluster shows a very high mean. This is because there were only 17 individuals there out of which 1 person had been assigned 45 secondary infections. Similarly, the “Pharmaceutical Company Nanjangud” had one person among 22 who was assigned 25 secondary infections. In Figure 9 we observe that the mean number of offsprings of parents is higher than 1.5. The four clusters where source of infection of the parents is not known are “Containment Zone”, “SARI”, “ILI” and“Unknown”clusters. Within these, it can be observed that mean of secondary infections in the first generation is highest for “SARI”, followed by “ILI”, then “Unknown” and “Containment Zone”. This is because the first generation in “SARI”/“ILI” cluster are those with a history of SARI/ILI, displaying symptoms, and hence have a high viral load of the infection. Further, one could infer that severe restrictions by definition in the “Containtment Zone” are proving effective with mean of secondary infections across all generations being less that one. Finally, the parents in the “Unknown” cluster presumably consist of patients, who at the time of testing, had mild symptoms or were asymptomatic patients (being part of random testing conducted routinely). If this is definitive, then one could conclude that the effective mean reproduction number for patients in this category is given by that of the “Unknown” cluster.
- **Variation with Age**We consider the age distribution across “The 8 clusters” in Figure 10. It is seen that the distribution of the coronavirus patients has a higher fraction of the patients in the ages 25 and above, whereas not too many in the range 0 *−* 25, when compared to the actual demographic distribution in Karnataka. A possible reason for this might be that the many cases were mainly restricted to travel of working professionals. The state also took steps quite early on to lock down schools and universities to prevent the younger segment of the population from being affected. The patients in the age group of 0 *−* 15 are either primary or secondary contacts of someone in their respective cluster. In Figure 11, we consider a heat map of ages across “The 8 clusters”. Patients below the age of 10 and those whose ages are greater than 90 have very very low mean of number of secondary infections caused. Most secondary infections are caused by middle aged people. The most socially active ones. For both “SARI” and “ILI” the age group 70 *−* 90 have higher means. This could be because of care takers and close family contracting the infection before the patient tested positive. The “TJ Congregation” has a higher mean across all groups from 10 *−* 80 and the “Pharmaceutical Company Nanjangud” as well has similar features. This is perhaps due to the fact that “TJ Congregation” cluster arose from a meeting in Delhi and the “Pharmaceutical Company Nanjangud” consisted solely of company employees and their Before 26^th^ June, 95% of the cases and 59% of the deaths occured in individuals less than 65 years old. The overall deceased rate is 2.414%. Among the patients in Karnataka who were deceased, 66% did so before they tested positive. Among the deceased patients who tested positive while hospitalized, the median number of days before they passed away was 3. The highest number of days a patient was treated before passing away was 36. From the detailed information on deceased patientes, it is also known that around 70% of them have comorbidities. We have computed the recovery rates and deceased rates for “The 8 clusters” in Table 7. We also plot the days to recover (in Figure 12a) and days to decease (in Figure 12b) among patients who tested positive before 26th June belonging to “The 8 clusters”. It is also seen that many patients who have passed away, do so on Day 0. This is because their samples, which result in positives, were sent for testing after their passing. It can be observed that bulk of the deceased patients are between 45 *−* 75 years. There does not seem to be any observable correlation with regard to days to recover and age. We caution against making significant inferences from this graph as the ‘recovery’ policy has changed with time (See for e.g. 1^st^ April and 8^th^ May Guidelines).
- **Variation across Districts**In Figure 13 we have plotted a heat map of the mean number of secondary infections in each week for the different districs. This provides a framework for the time evolution of the reproduction number across districts as done in comparmental models. Most districts in Karnataka started having their COVID-19 cases quite late, during early May. Bangalore-Rural, Bellary, Davangere, Dharwad, Karwar, Kodagu, Tumkur and Udupi, have several weeks where no one tested positive, as earlier outbreaks were well contained. The Pharmaceutical in Nanjangud is in the Mysore district and the end of the outbreak is visible. In Davangere District in week 27-Apr and 3-May, has a large mean becaused of a patient who was infected on 29-Apr and had been assigned 30 secondary infections and one on 30-Apr who was assigned 18 secondary infections. This is typical when there is a large mean. Most of the cases in the districts have low mean number of secondary infections in May. This was mainly due to the fact that those tested positive in this month had migrated from other states and caused very few recorded secondary infections.

**Table 3:** The above table is an extension of Table 2. The column ‘*k*’ contains the Maximum Likelihood estimates for the dispersion parameter, *k*, and the column ‘*p*-value’ contains *p*-value from the *χ*^2^ goodness of fit test of the Negative Binomial Distribution fitted to the Data. (see Sections B and C for details.)

| Name of Cluster | Size | Maximum | $R_{\text{eff}}$ | Variance | $k$ | p-value |
| --- | --- | --- | --- | --- | --- | --- |
| Containment Zones | 293 | 7 | 0.3447 | 1.199 | 0.09345 | 0.5966 |
| From the Southern States | 286 | 7 | 0.2168 | 0.6897 | 0.08424 | 0.5409 |
| ILI | 1398 | 30 | 0.3369 | 2.355 | 0.06428 | 0.736 |
| Others | 707 | 51 | 0.5191 | 6.502 | 0.09214 | 0.8318 |
| Pharmaceutical Company | 73 | 24 | 0.726 | 8.757 | 0.1839 | 0.002671 |
| SARI | 746 | 45 | 0.6743 | 8.521 | 0.08023 | 0.4698 |
| TJ Congregation | 97 | 15 | 0.7732 | 3.823 | 0.2138 | 0.1138 |
| Unknown | 1295 | 27 | 0.2625 | 1.637 | 0.05792 | 0.1225 |

**Figure 6:**
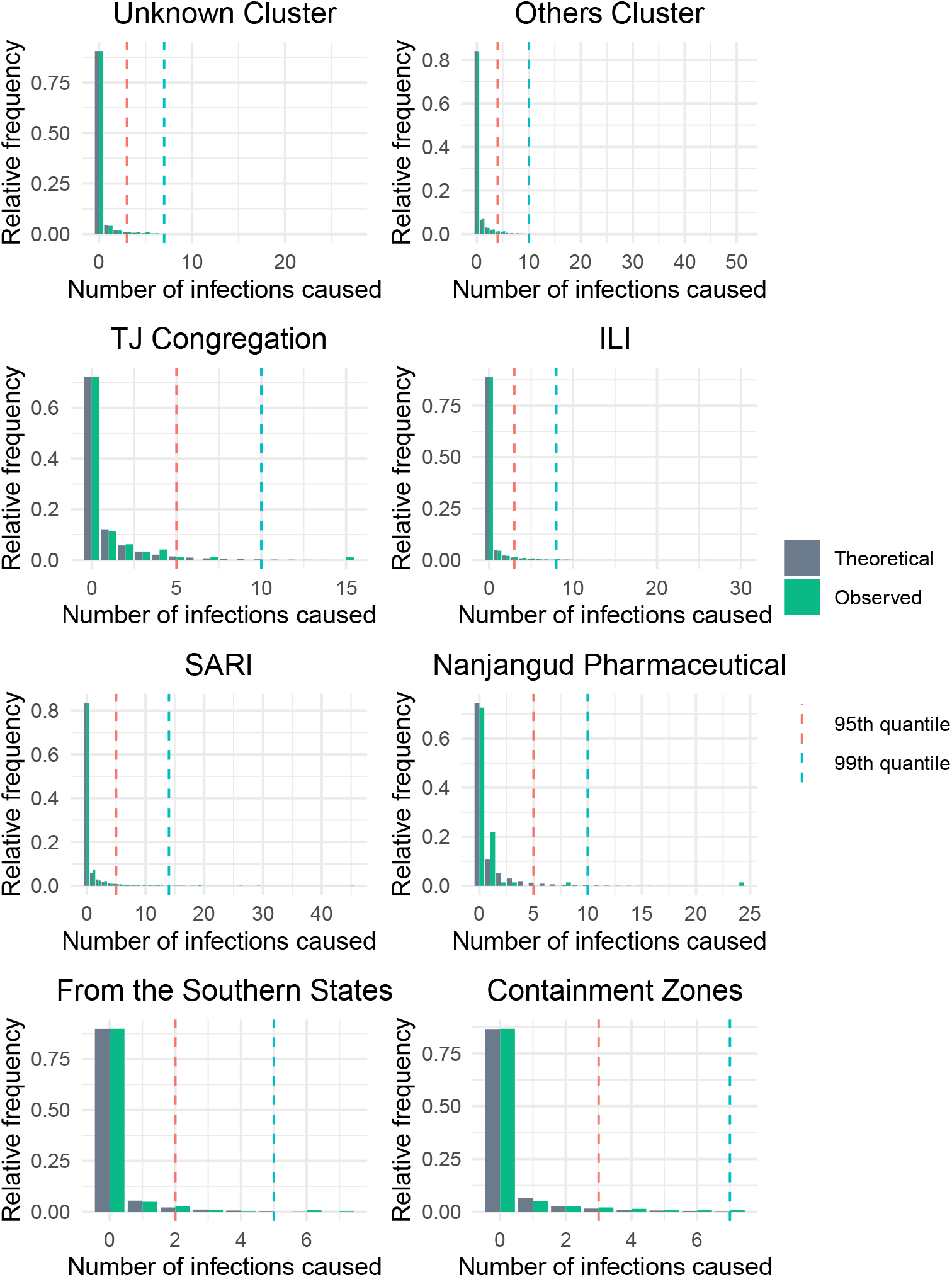
The above graph plots for each cluster the observed relative frequencies of the offspring distribution along with the theoretical negative binomial probabilities. The green bars represent the observed relative frequencies and the grey bars represent the probabilities as calculated from the negative binomial distribution. The graph also has the 95th and 99th quantiles of the calculated negative binomial distribution marked in red and blue respectively. One can use this to compute the probability of super-spreading events.

**Table 4:**
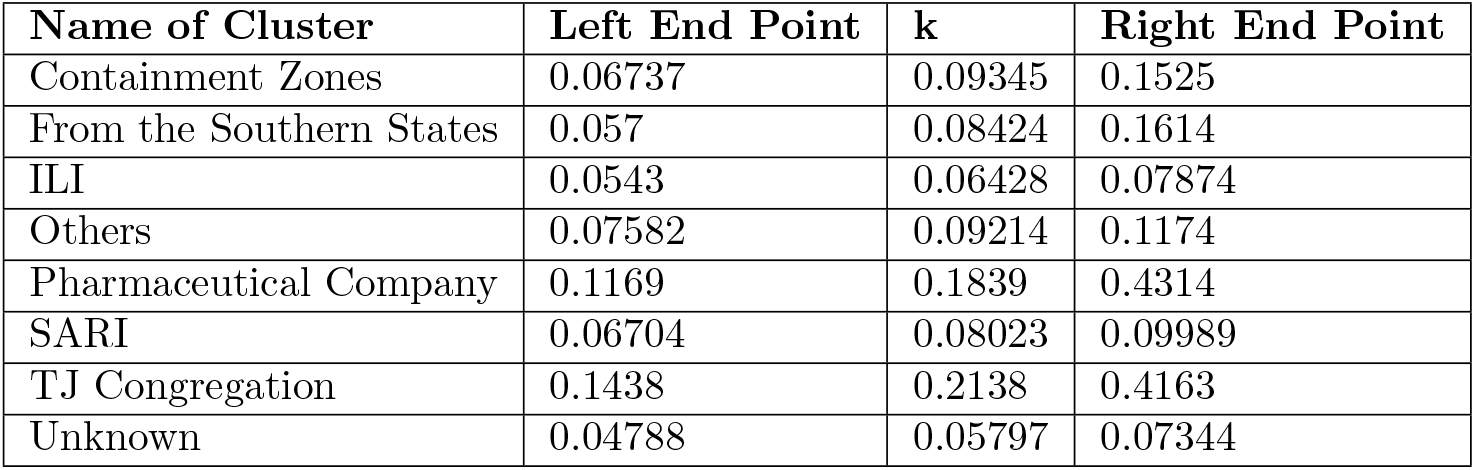
The above table provides 95% confidence intervals for *k*. See Section D for methodology used to calculate them.

**Figure 7:**
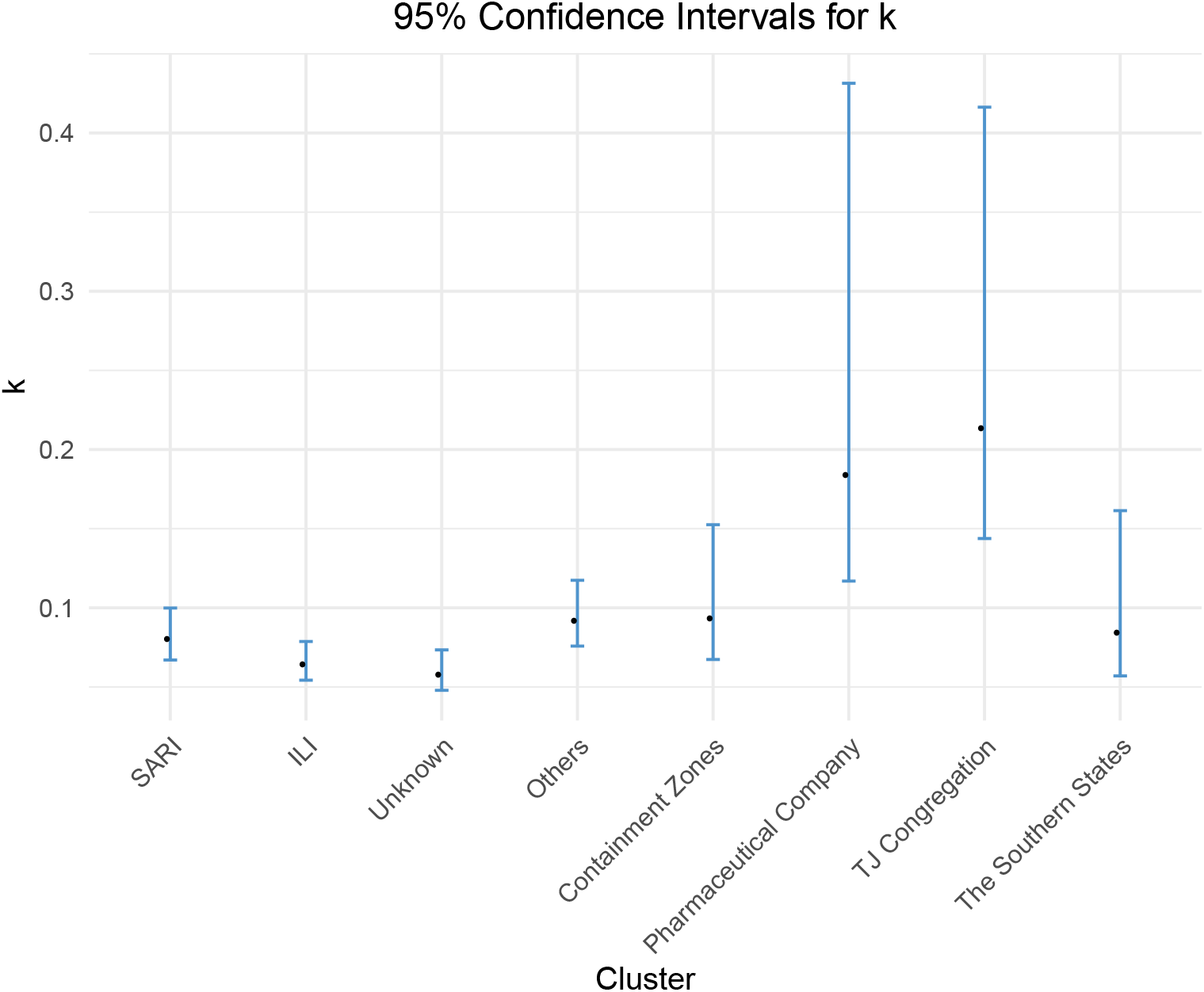
The above plot contains the calculated values of the dispersion parameter, *k*, along with its 95% confidence intervals for each of “The 8 clusters” in observation.

**Table 5:** The above table computes the 90^th^, 95^th^ and 99^th^ quantiles of the Poisson(*R*_eff_) distribution. The quantiles will define 90^th^, 95^th^ and 99^th^-Super-spreading events respectively.

| Name of Cluster | $R_{\text{eff}}$ | 90<sup>th</sup>-percentile | 95<sup>th</sup>-percentile | 99<sup>th</sup>-percentile |
| --- | --- | --- | --- | --- |
| Containment Zones | 0.3447 | 1 | 1 | 2 |
| From the Southern States | 0.2168 | 1 | 1 | 2 |
| ILI | 0.3369 | 1 | 1 | 2 |
| Others | 0.5191 | 1 | 2 | 3 |
| Pharmaceutical Company | 0.726 | 2 | 2 | 3 |
| SARI | 0.6743 | 2 | 2 | 3 |
| TJ Congregation | 0.7732 | 2 | 2 | 3 |
| Unknown | 0.2625 | 1 | 1 | 2 |

**Figure 8:**
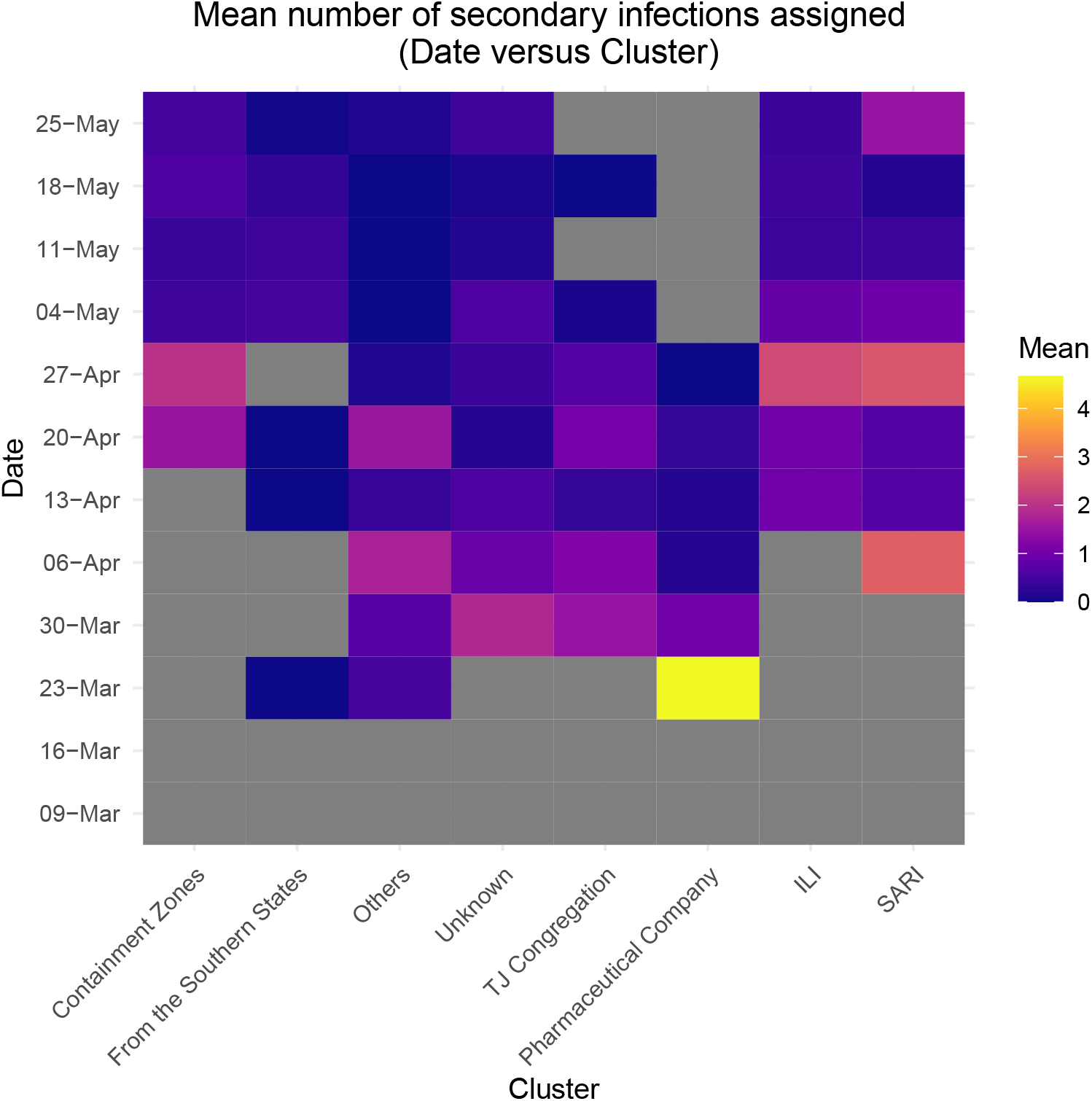
In the above plot, each tile represents a cluster and a week starting at the date given on the left. The color of each tile represents the mean number of infections caused by the patients infected during that week belonging to the cluster. A grey tile means there are no individuals in that cluster who were infected in that particular week. It can be observed in the plot how *R*_eff_ for each of the clusters varies over time. This can be compared to *R*_t_ that is observed using compartmental models as in [6].

**Table 6:** The above table considers the different generations of infections as seen in Karnataka for “The 8 clusters”. For each generation, the table contains the number of individuals in that generation, the number of patients causing zero secondary infections, the maximum number of infections caused by an individual in that generation and the mean number of infections caused by an individual in that generation.

| Name of Cluster | Size | Zero | Maximum | Mean |
| --- | --- | --- | --- | --- |
| Parent | 2925 | 2528 | 30 | 0.4499 |
| Child | 1314 | 1176 | 51 | 0.3067 |
| Grandchild | 403 | 344 | 14 | 0.3524 |
| Great Grandchild | 142 | 112 | 45 | 0.7042 |
| Great Great Grandchild | 100 | 94 | 5 | 0.11 |

**Figure 9:**
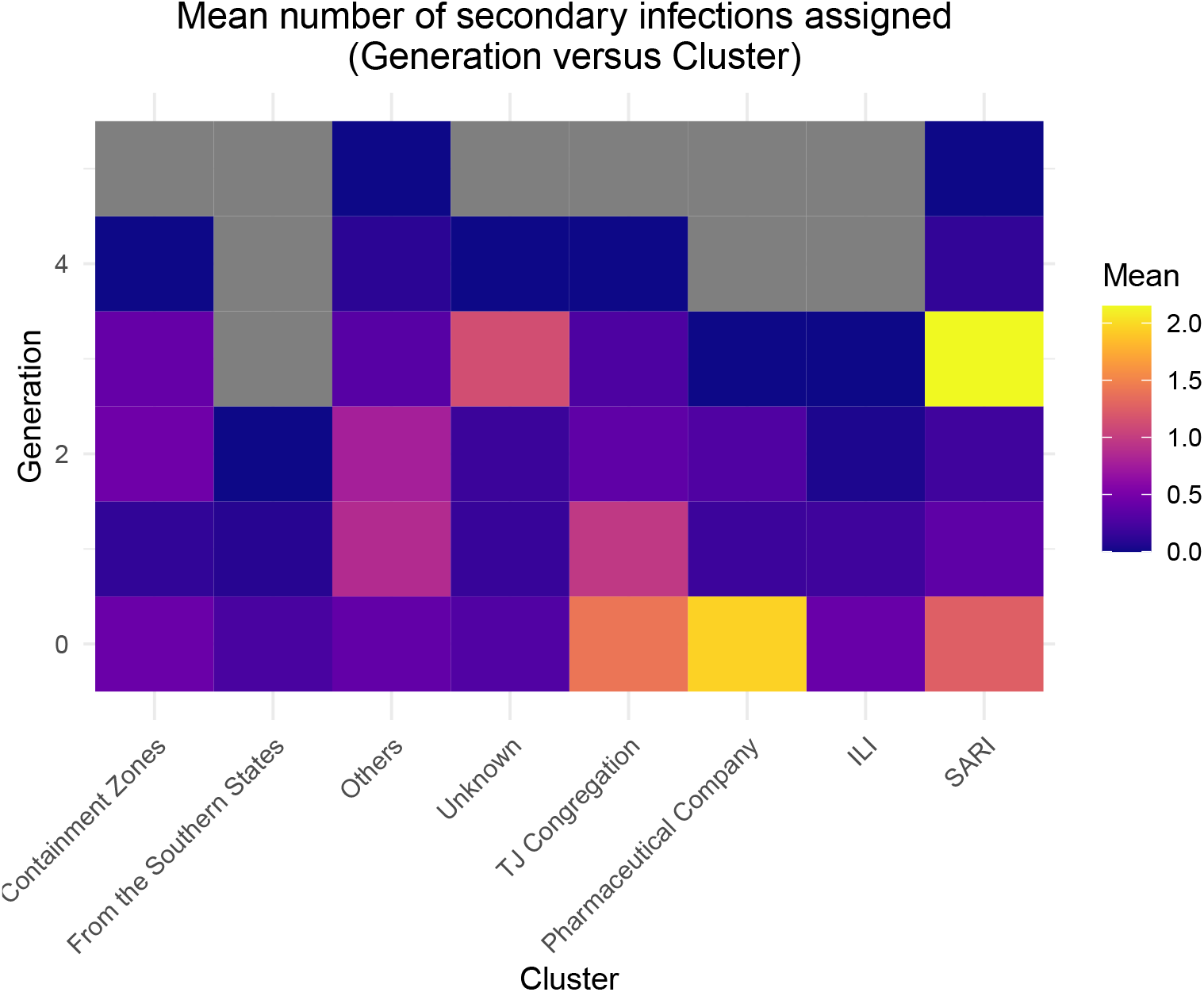
Generation 0 refers to those patients who were the first in their respective clusters to be identified as infected. They may have had either some form of travel history or displayed certain symptoms. Generation *x* + 1 represents the individuals who were infected by Generation *x*. The color of each tile represents the mean number of infections caused by each individual belonging to a particular cluster and generation.

**Figure 10:**
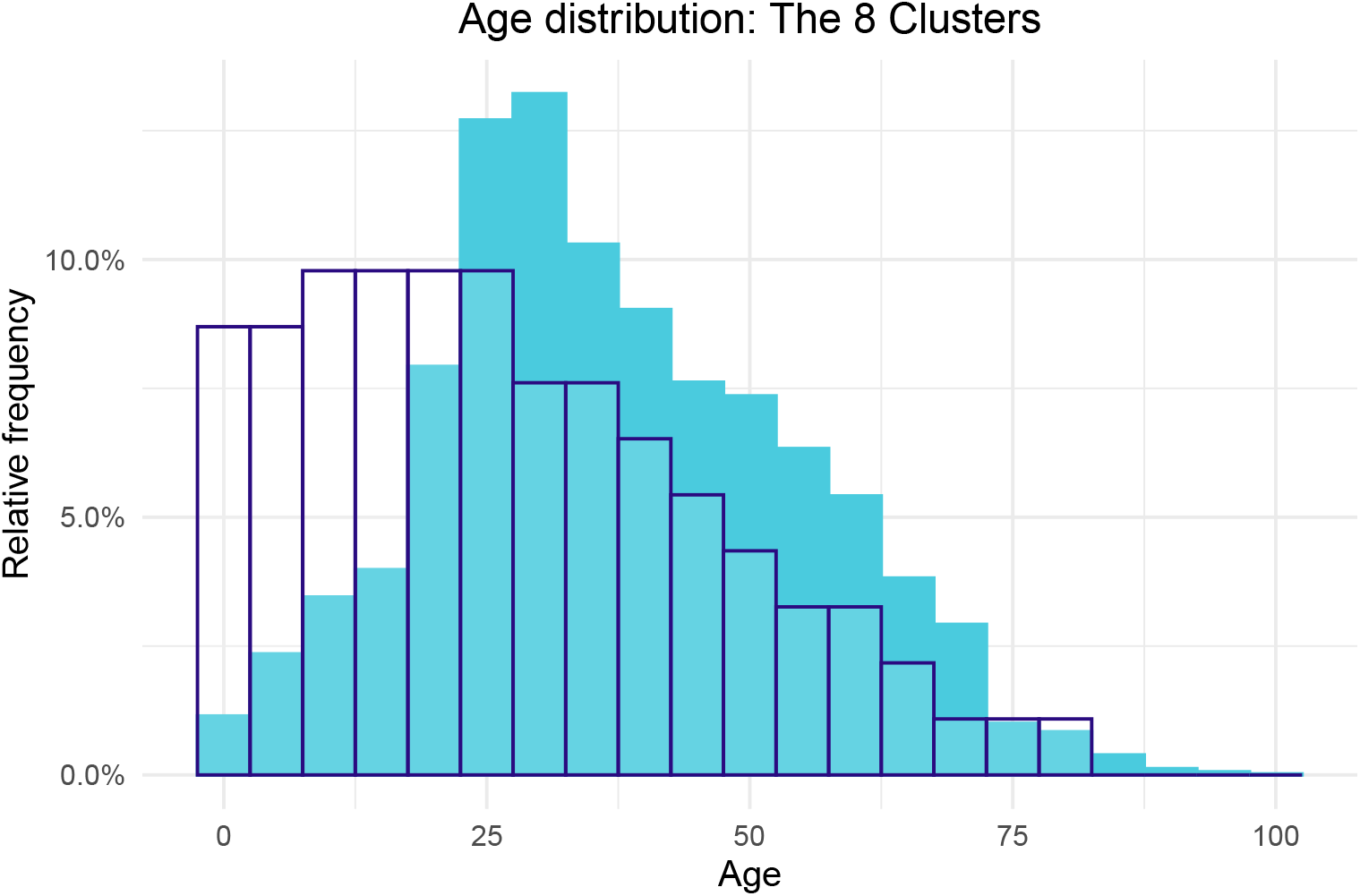
For the patients belonging to “The 8 clusters” in study, the above histogram contains the distribution of their ages. The filled in light blue color indicates the distribution of the coronavirus patients in Karnataka. The navy outline in both graphs represents the Age distribution of the entire population of Karnataka for reference (taken from the 2011 census).

**Figure 11:**
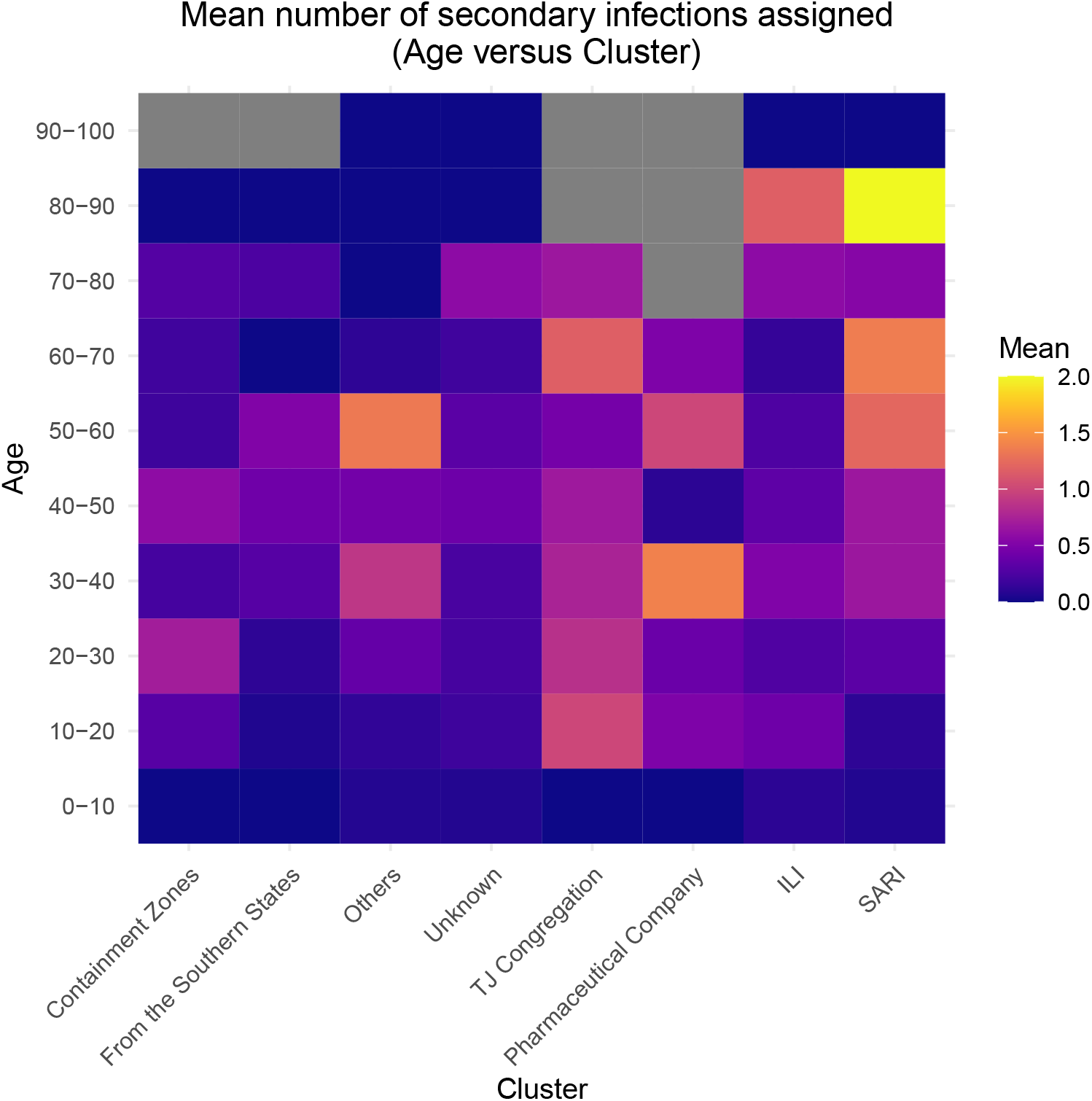
Each tile in the above plot represents a cluster and an age bracket. The color represents the mean number of offsprings for those patients falling into the respective age bracket and cluster. The grey tiles represent the lack of patients falling in that particular demographic.

**Table 7:** The above table contains information on “The 8 clusters” for those patients who tested positive before 26th June. For each cluster, the recovery and deceased rate has been listed in the table.

| Name of Cluster | Recovery Rate | Deceased Rate |
| --- | --- | --- |
| Containment Zones | 96.06 | 3.93 |
| From the Southern States | 95.91 | 4.08 |
| ILI | 91.21 | 8.78 |
| Others | 98.06 | 1.93 |
| Pharmaceutical Company | 100 | 0 |
| SARI | 82.68 | 17.31 |
| TJ Congregation | 98.75 | 1.25 |
| Unknown | 93.08 | 6.91 |

**Figure 12:**
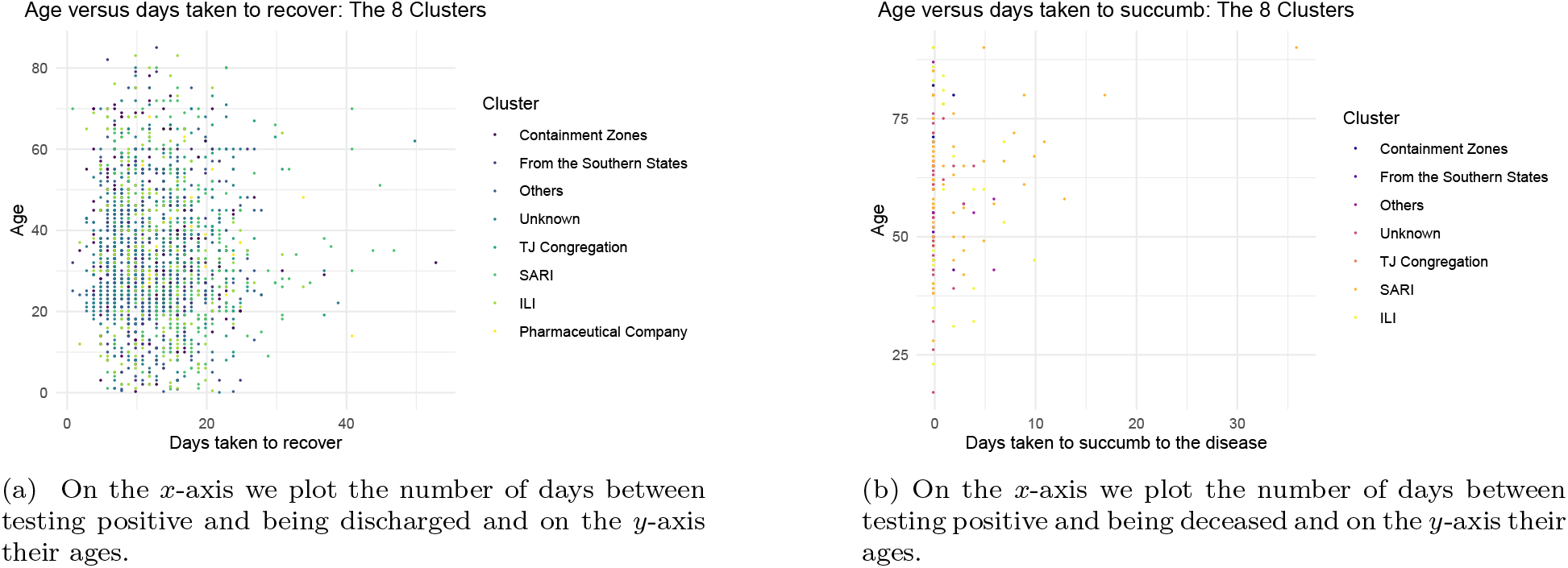
The above scatter plot contains information about “The 8 clusters”. The different points have been colored based on the clusters.

**Figure 13:**
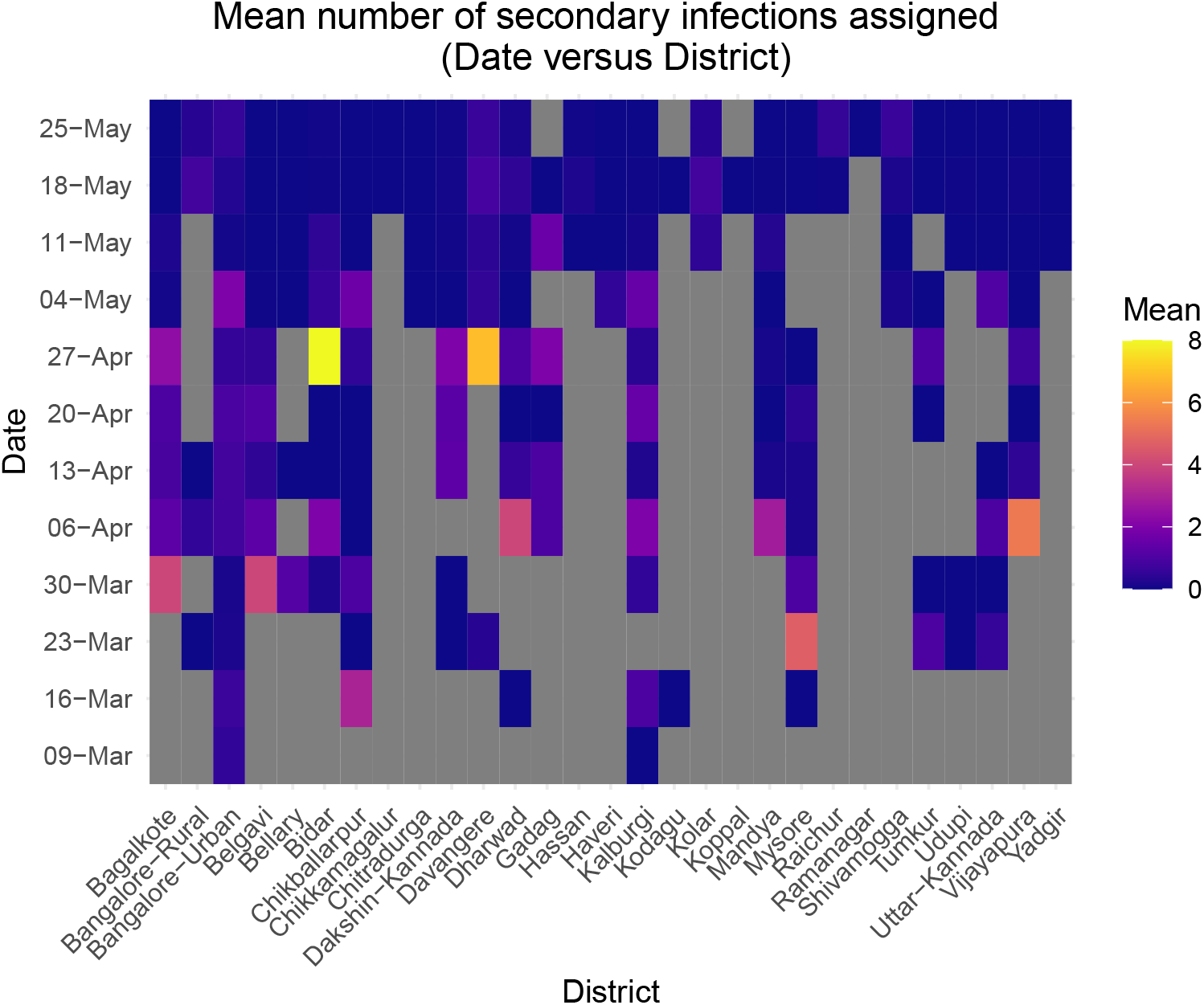
To see the varying spread of the disease in different districts, we have plotted a heat map of the mean number of offsprings in each week for the different districts. Every tick on the *y*-axis represents a week starting at the given date. The *x*-axis represents the different districts. Each tile represents the mean number of infections caused by those patients.

### Cases due to Migration in Phase 3,4 and Unlockdown 1.0

As mentioned earlier, we had omitted two clusters from analysis, namely: From Maharashtra and From Middle East. Phase 3 and 4 of the lockdown, along with Unlockdown 1.0 in June loosened restrictions on Domestic Travel and International travel. The state saw a large influx of infected individuals from within India and abroad. During Phase-3 of the lockdown, the “From Maharashtra” cluster saw the most growth and dominated the test positive counts by a significant margin. The ‘From Maharashtra’ cluster accounted for approximately 52.5% of cases in the stipulated period. The“From Middle East” cluster seems to have two phases. The first, before the lockdown was enforced during which international travel were suspended. The second, more recent, due to the repatriation flights from the region. We provide the Maximum Likelibood estimators for *R*_eff_ and *k*, along with the summary of them in Table 8. During this period domestic and international travellers were quarrantined/tested on arrival. To make any meaningful inferences using reproduction numbers and dispersion one would have take into account a more detailed tracing history procedure from their origin of travel.

**Table 8:** For the two clusters “From Maharashtra” and “From Middle East” an analysis similar to that of Table 3 has been done.

| Name of Cluster | Size | Maximum | $R_{\text{eff}}$ | Variance | $k$ | p-value |
| --- | --- | --- | --- | --- | --- | --- |
| From Maharashtra | 5486 | 30 | 0.03992 | 0.3804 | 0.007059 | 0.5942 |
| From Middle East | 409 | 6 | 0.02934 | 0.1168 | 0.007965 | 0.3165 |

To understand cases due to Migration (6871 out of 10391) in this period we reorganized our clusters (See Table 1) into four groups: the first group being referred to as the “Foreign group” (379 cases), consists of patients who belong to the “From Middle East”, “From United Kingdom” and “From the rest of Europe” cluster as well as a few cases which originated from Nepal, Indonesia, Philippines and Malaysia; the second group being patients who belong to “From Gujarat”, “From Rajasthan”, “From the Southern States” (Kerala, Tamil Nadu, Telangana and Andhra Pradesh) and “Others cluster” who had traveled to Delhi have been combined to form the “Inter-State Travel” group (582 cases); the third group being patients who belong to “From Maharashtra” cluster (5481 cases) and will be denoted by the same name; and finally the fourth group are patients who belong to “Others” cluster whose testing positive is attributed to inter-district travel within Karnataka, which we have called the “Inter-District Travel” group (429 cases).

- **Variation across Districts**The “From Maharashtra” group (or cluster) affected the districts of Kalaburagi (1113 cases), Udupi (1020 cases) and Yadgir (891 cases) the most. Bangalore-Urban received only 85 cases from Maharashtra (See Figure 16). The “Inter-State Travel” group affected Bangalore-Urban and Mysore the most, though with absolute numbers being very low at 99 and 46 cases respectively (See Figure 17). The“Foreign group”contributed 379 cases (via airports in Bangalore and Mangalore) with 76% were detected in the Dakshina Kannada District (See Figure 18) and 43 of them were assigned to Bangalore-Urban district. The “Inter-District Travel” group affected Ballari the most, with 214 cases. Mysore and Bangalore-Urban ranked next, but their absolute counts were quite low at 49 and 30 respectively (See Figure 19). As seen from above, overall, Kalaburagi, Udupi and Yadgir were the worst affected districts. The three districts together received 45.2% of all infections due to Migration and Bangalore-urban received 257 cases due to migration (See Figure 20).
- **Migration versus Total**In Table 9 we compute the percentage of cases due to the migration group across districts during this period. We observe that Dakshina Kannada (84%), Kalaburagi (89.8%), Mandya (95.3%), Raichur (90%), Udupi (94.3%) and Yadgir(98%) had very proportion of their total cases due to migration group. In contrast, in Bangalore-Urban migration accounted for 14.3% of the total cases (See Figure 21).
- **Variation in Age, Recovery and Deceased**The histogram of age distribution due to migration cases shows that the distribution is concentrated around 20–40 years as seen in Figure 14. There are also a higher proprtion of cases having 0–20 age as compared to the histogram of all cases and a lesser proportion of elderly people as seen in Figure 10 (which has the age distribution for the infected individuals belonging the the eight clusters in study earlier), indicating that most of the migrating individuals were families. This is probably because more children migrated along with parents, but very few elderly people did. Out of the 6871 migration cases, only 25 people have succumbed to the disease (as seen till 21st July). This is perhaps due to the fact that the elderly were in fewer proportion than in the 8 clusters that we analyses earlier. There were no casualities in the “Foreign group”. All but one person who passed away were more than 40 years of age as seen in Figure 15b. There was also a high recovery rate with 6657 people recovering (as seen till 21st July). Most people recovered within 20 days of testing positive as seen in Figure 15a. Again we caution against making significant inferences from this graph as the “recovery” policy has changed with time.

**Figure 14:**
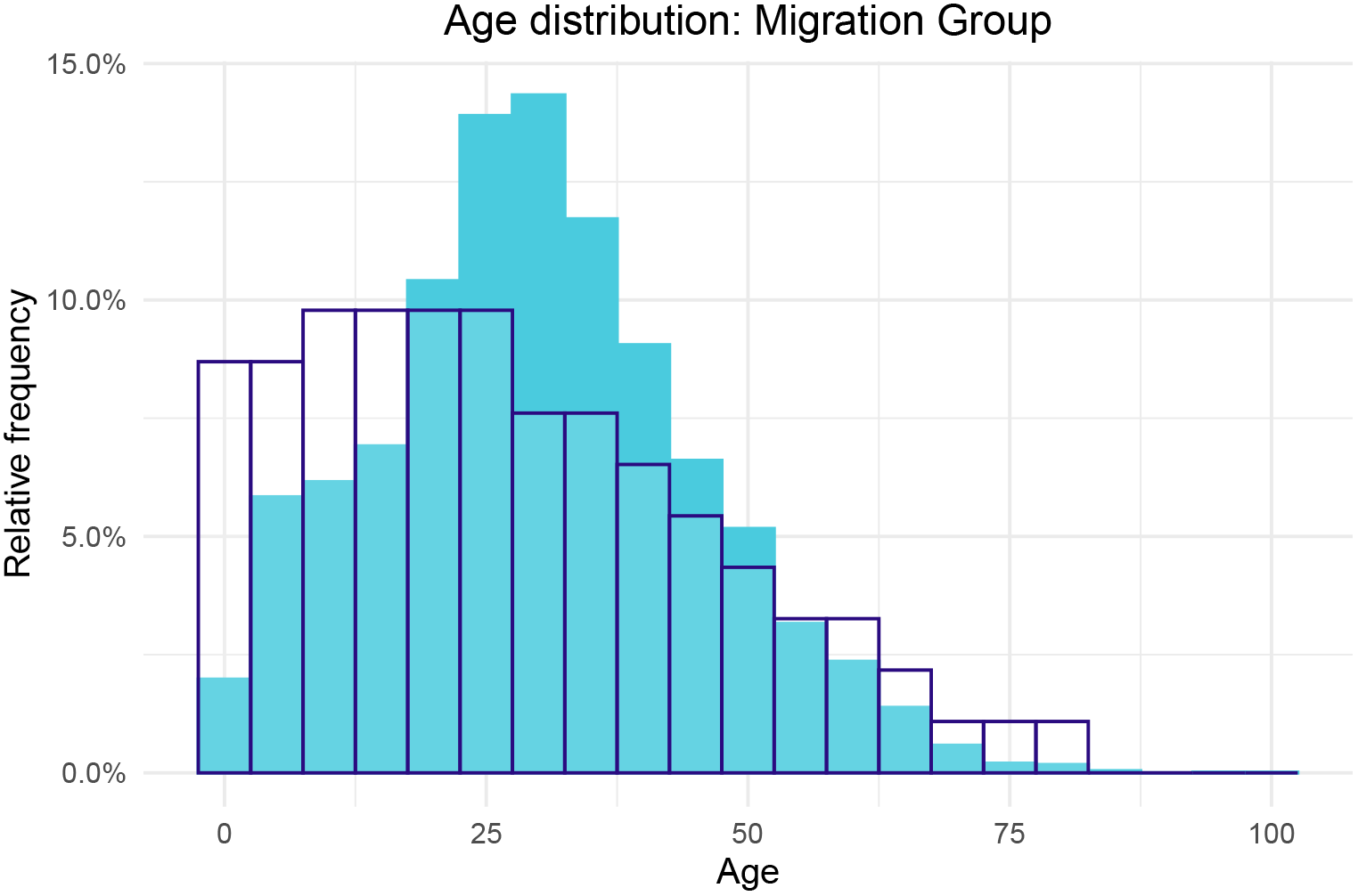
The above graph is a histogram for the individuals who belong to the Migration group. The filled in light blue color indicates the distribution of the patients belonging to the Migration group. The navy outline in both graphs represents the Age distribution of the entire population of Karnataka for reference (taken from the 2011 census).

**Figure 15:**
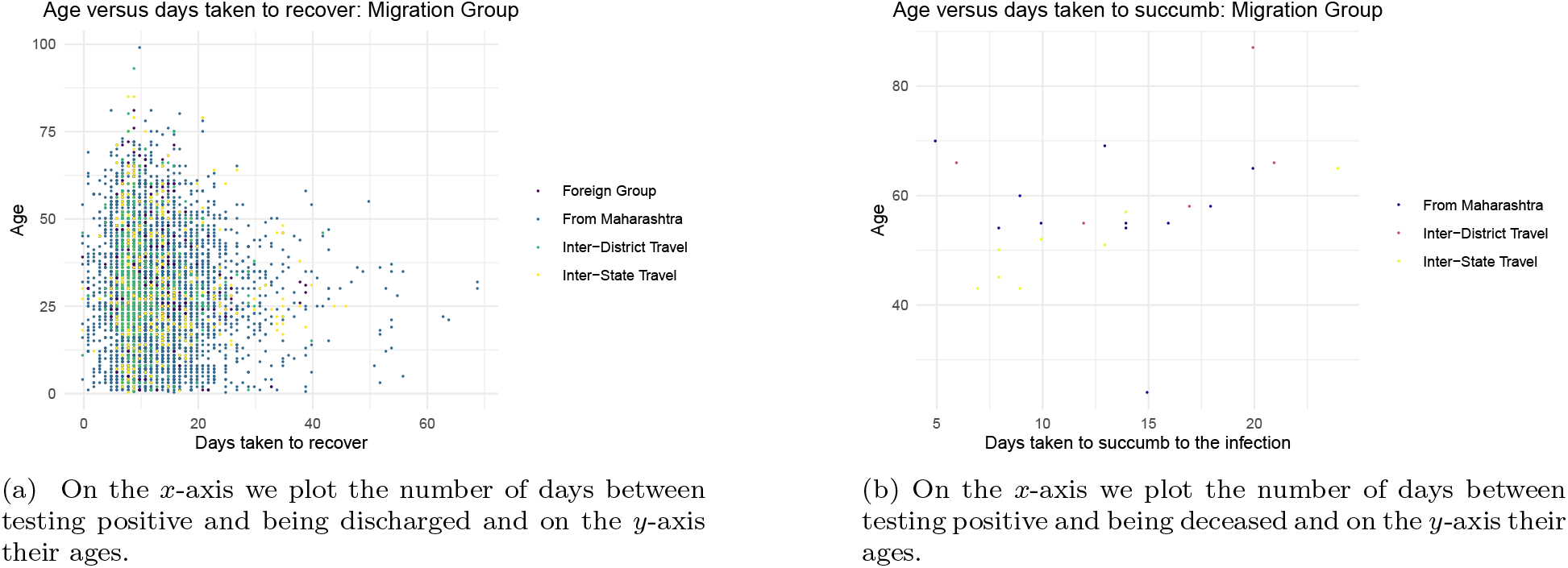
The above scatter plot contains information on the individuals belonging to the Migration group. The colors represent the different groups.

**Figure 16:**
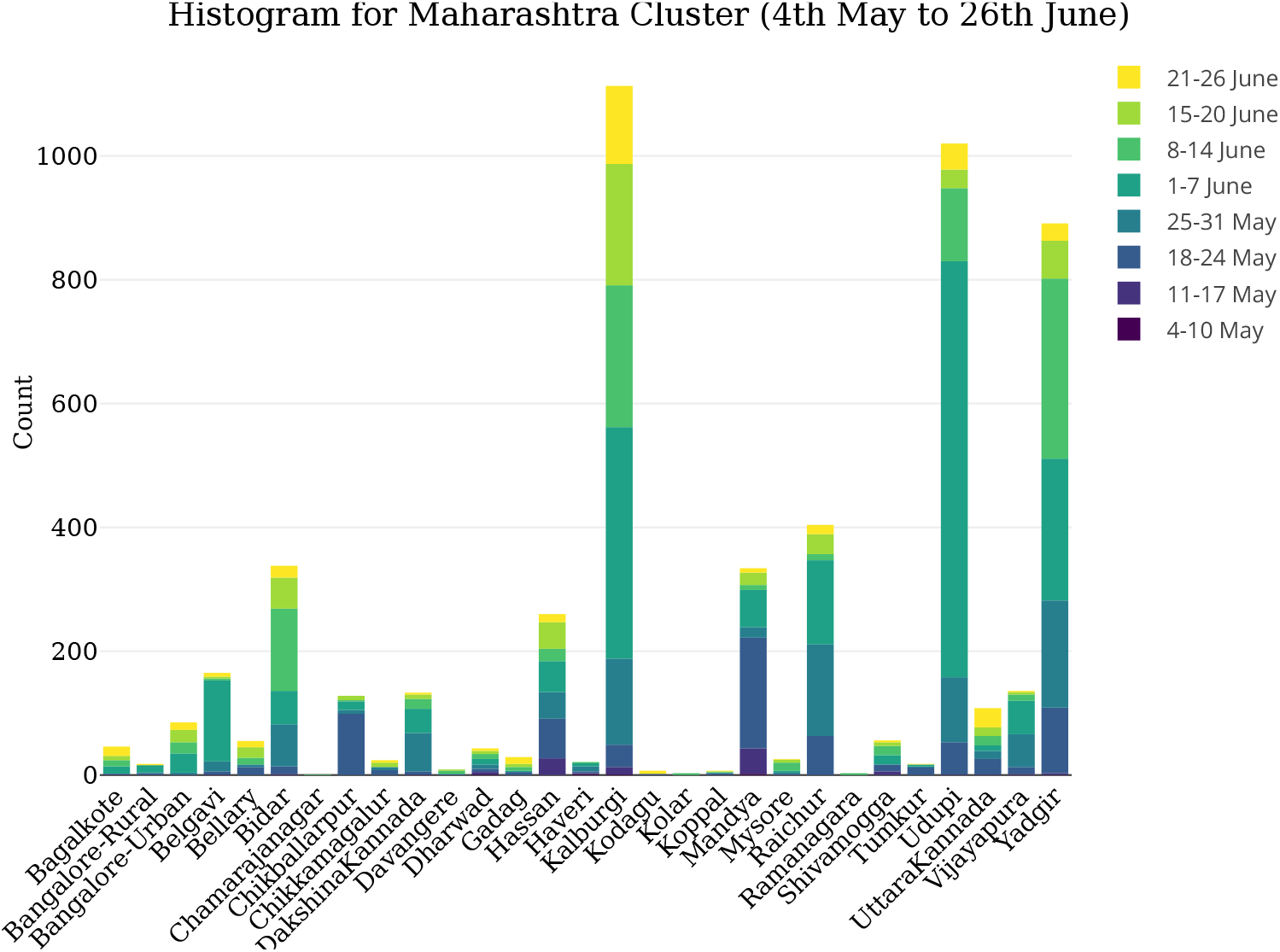
This figure represents the “From Maharashtra” cluster in the migration period (4^th^ May to 26^th^ June). The cases are separated district-wise on the x-axis and the bar height is the number of cases “From Maharashtra” cluster in the given district. The bar has been divided into colors according to the time period mentioned in the legend. In this graph, there are a total of 5481 infections, out of which Kalaburagi, Udupi and Yadgir were most affected with 1113, 1020 and 891 cases respectively. Bangalore-Urban had only 85 (around 1.5%) cases

**Figure 17:**
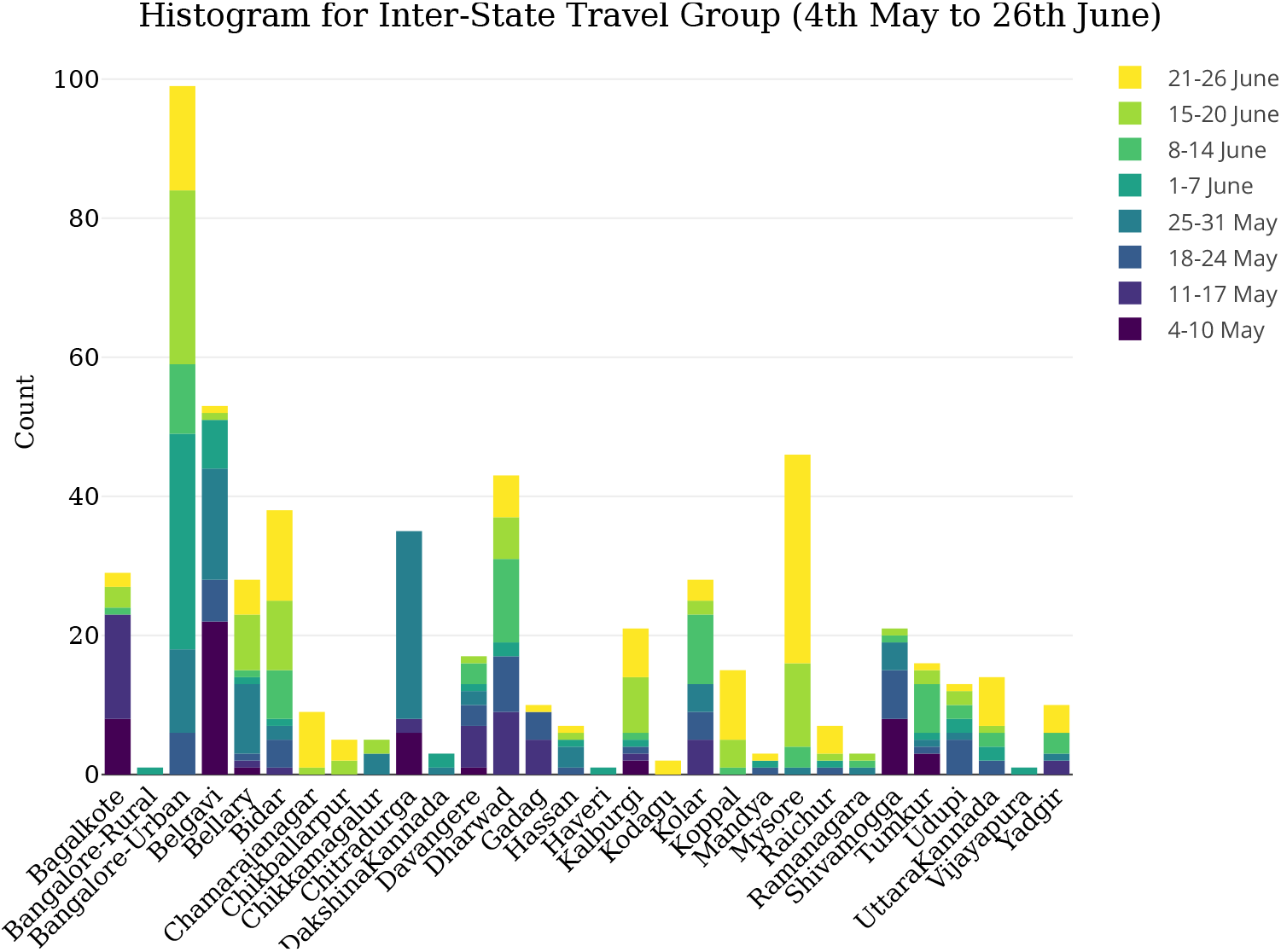
This graph represents the migration cases due to the “Inter-State Travel” group. Please note by our definition this excludes the cases “From Maharashtra”. There are a total of 582 cases. 99 cases were recorded in Bangalore-Urban, the most of any district. However, the absolute number is quite less

**Figure 18:**
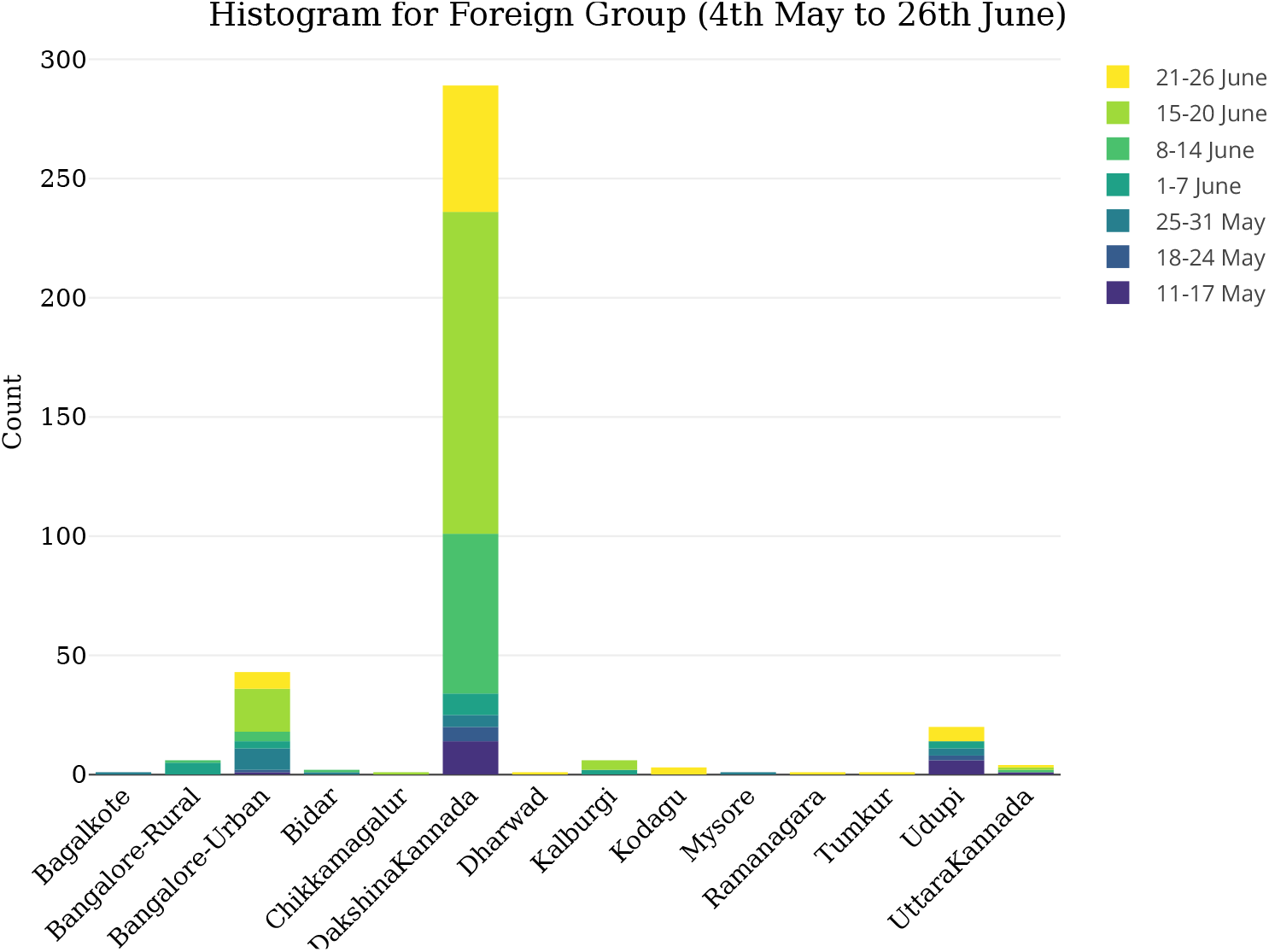
This graph represents the migration cases due to the “Foreign group”. There are a total of 379 cases, of which 159 were in the period 15 *−* 20^th^ June. 289 cases were detected in Dakshin Kannada, mainly Mangalore. Only 43 cases were detected in Bangalore-Urban

**Figure 19:**
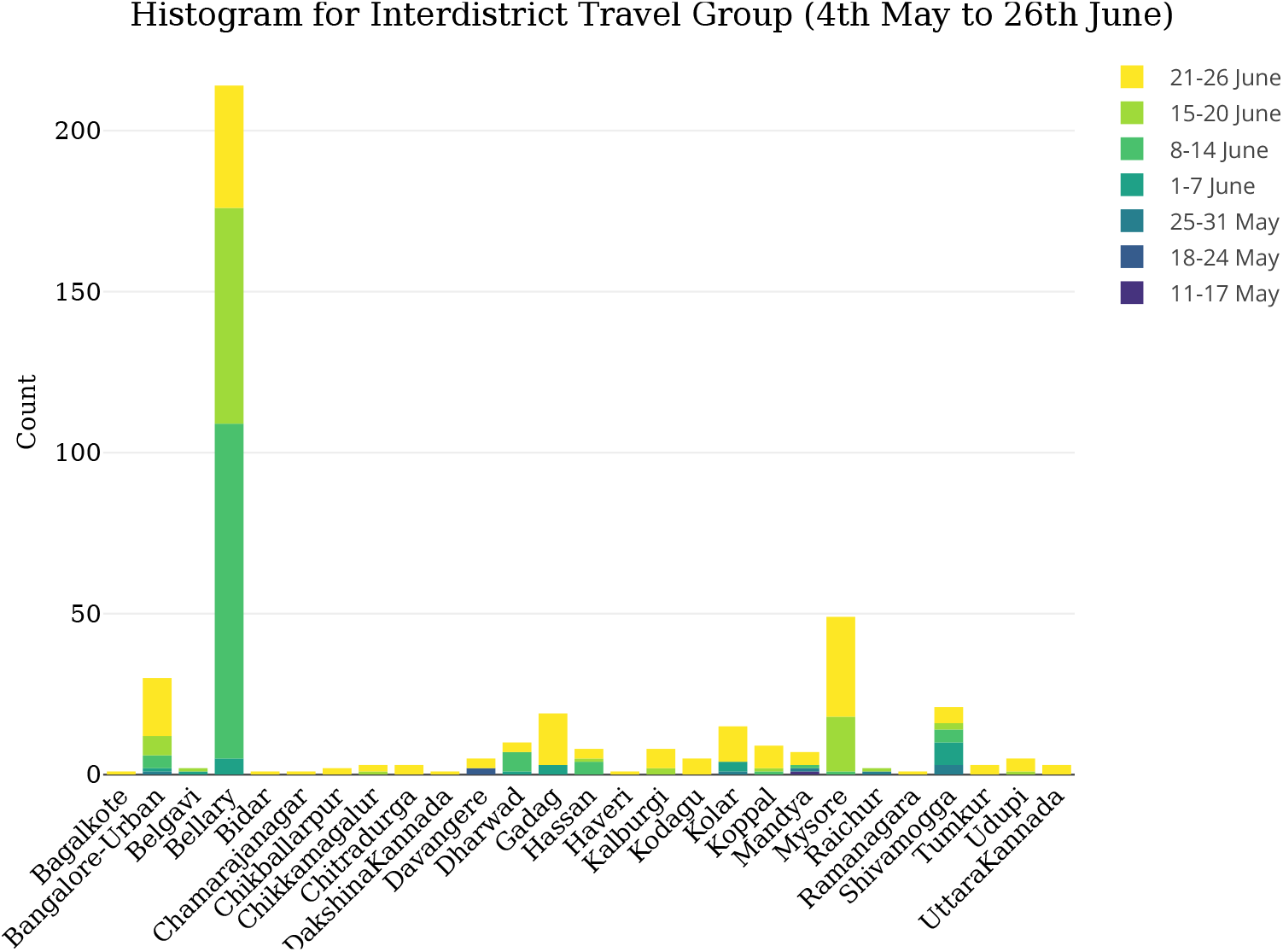
This graph represents the migration cases due to the “Inter-District Travel” group. There are a total of 429 cases. The most affected district is Ballari with 214 cases

**Figure 20:**
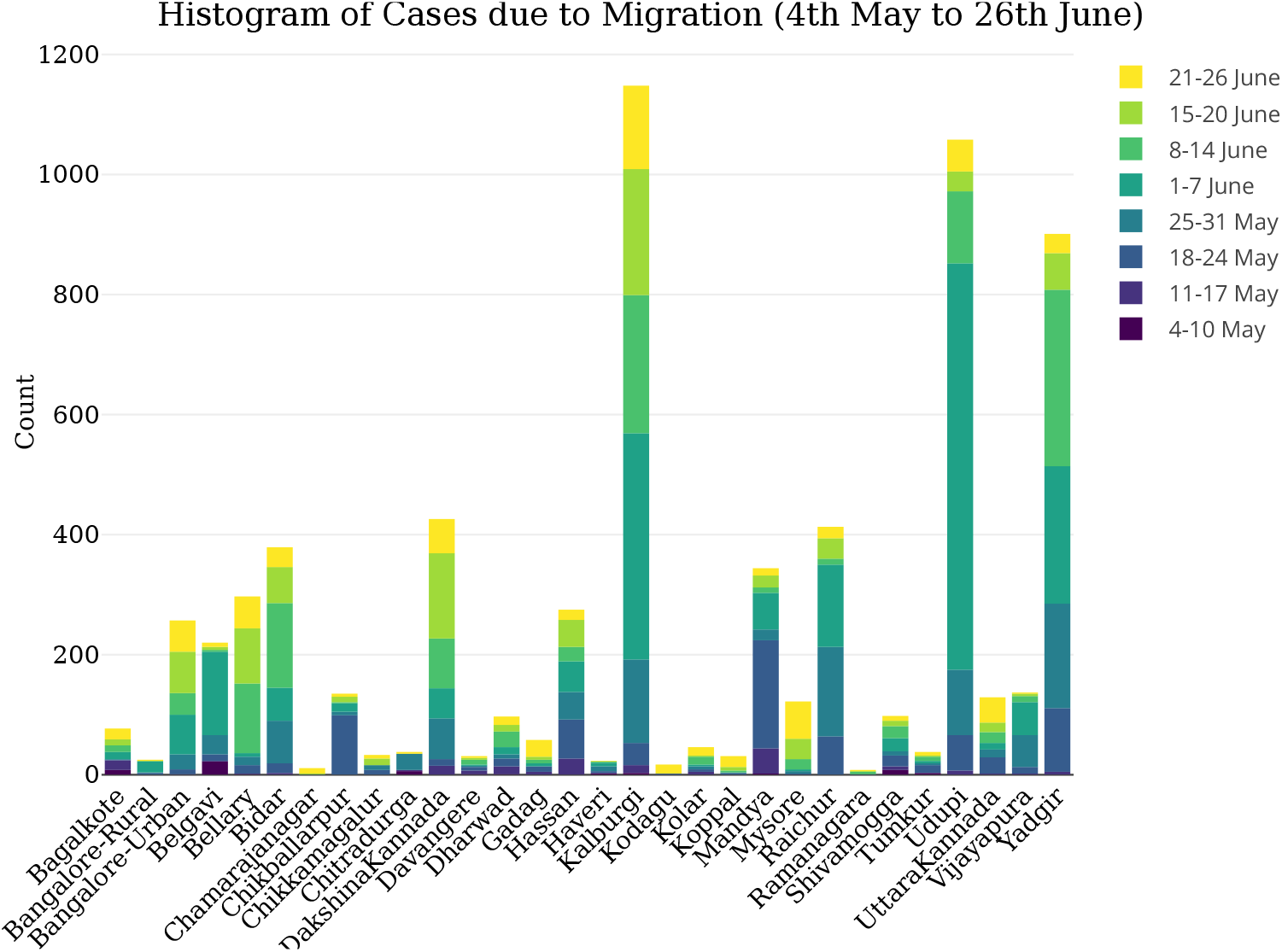
This graph represents all the migration cases from 4^th^ May to 26^th^ June. There are a total of 6871 cases. The worst-affected districts were Kalaburagi, Udupi and Yadgir with 1148, 1058 and 901 cases respectively. Together, they received 45.2% of all cases. Bangalore-Urban recieved only 257 cases.

**Table 9:**
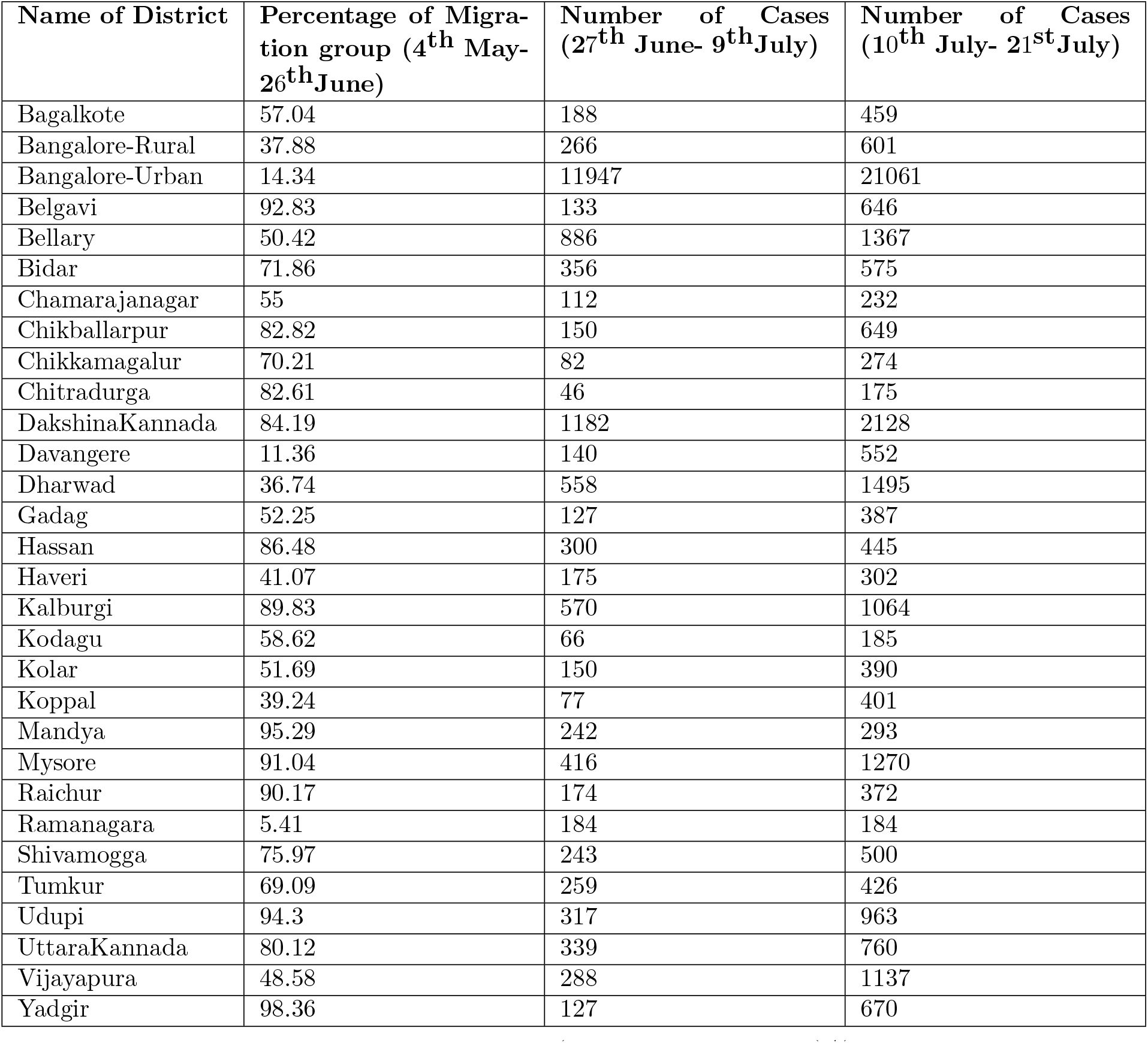
In this table, the first column represents (cases due to migration)/(total cases in district in the period 4^th^ May to 26^th^ June) as a percentage. The second and third column give absolute counts of cases in each district from 27^th^ June to 9^th^ July and from 10^th^ July to 21^st^ July respectively.

**Figure 21:**
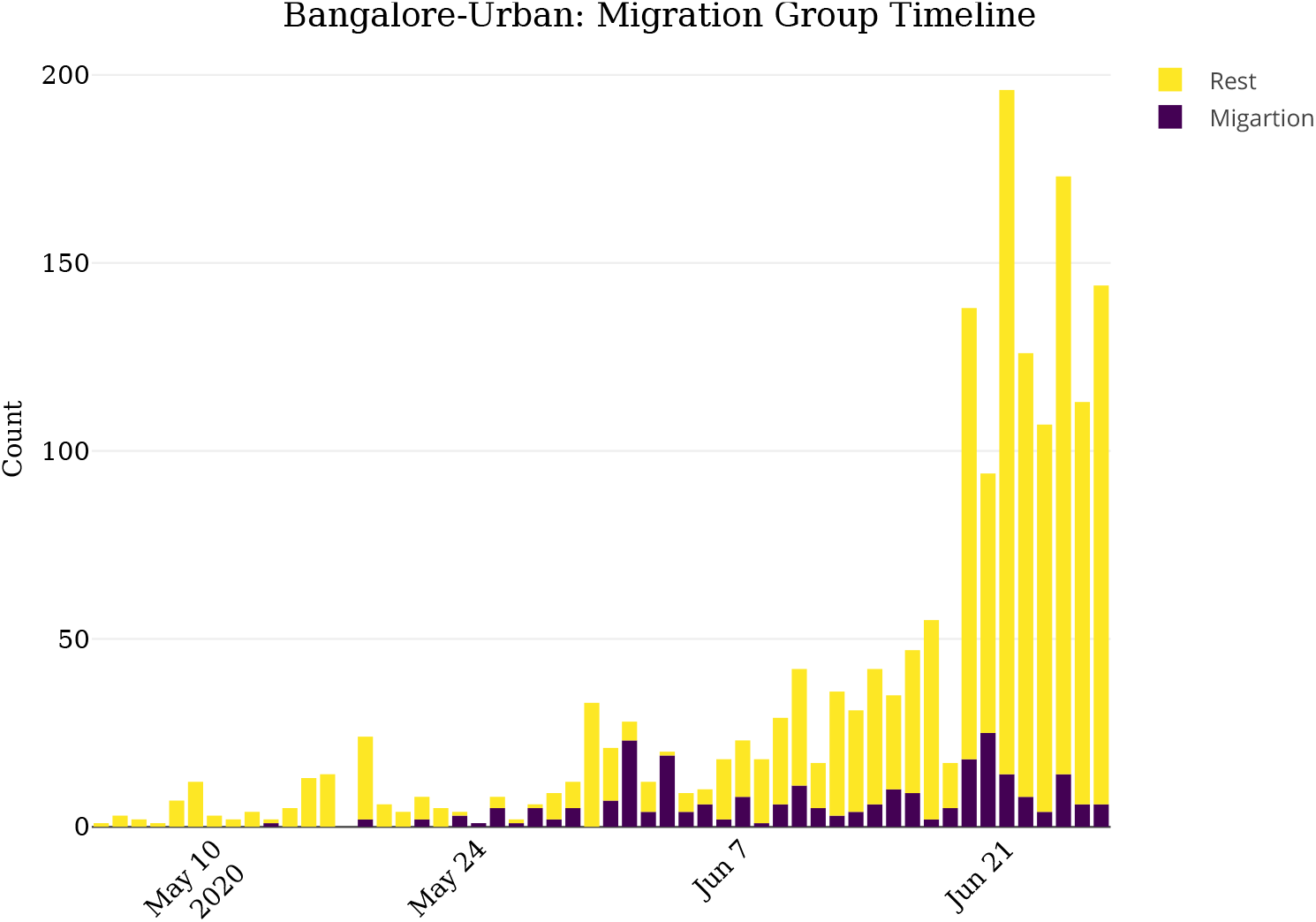
This graph represents the daily timeline of cases in Bangalore-Urban from 4th May to 26th June. The height of bars represent cases on that particular day. Each bar is divided into number of cases due to migration and cases due to other reasons. There were a total of 1792 cases out of which 257 were attributed to migration or contacts of those with a known history of migration.

### Surge in July

There was a sudden surge in cases in Karnataka after the migration period (4^th^ May to 26^th^ June). On 26^th^ June the total cases in the state stood at 11005, which doubled on 9^th^ July (31105 cases) and became four times on 21^st^ July (71068 cases). We will try to outline the possible reasons for this surge.

- **Contact under tracing/Unknown**From 27^th^ June to 29^th^ June the media bulletins did not provide any description for the patients who tested positive and from 30^th^ June onwards the description was not as detailed as before. Post 21^st^-July the media bulletins did not contain any individual information on those who tested positive (see Section 4 for details). A disporportionately large proportion of cases were designated as contact under tracing and thus falling in the “Unknown” cluster (see Figure 22a, Figure 22c). Thus it is not possible to proceed on a precise analysis as done for the 8 clusters.
- **ILI/SARI**From middle of June, the “ILI” cluster cases in Karnataka, have been increasing and there was a sharp rise in the first half of July (See Figure 24b and Figure 24a). They also formed a significant proporition of total cases (See Figure 23b and Figure 23a). In Bangalore-Urban district, the “ILI” cluster cases have been increasing since the middle of june, a sharp rise in the first half of July and also a significant proportion of total cases with over 50% on some days (See Figure 23c). The “SARI” cluster also shows an increase but the proportion fluctuates and is low around, 5%.
- **Variation across districts**Bangalore-Urban accounted for approximately 50% of the surge in July with the count being 1953 on 26^th^ June and rising to 34691 by 21^st^ July. In Kalaburagi, there were 1339 cases on 26^th^ June and 2973 cases on 21^st^ July. In Udupi and Yadgir, the cases doubled from 26^th^ June to 21^st^ July (Udupi- 1126 cases on 26^th^ June, 2406 cases on 21^st^ July; Yadgir- 916 cases on 26^th^ June, 1713 cases on 21^st^ July). We have plotted the timelines of all the districts in Figure 27 andFigure 28.

**Figure 22:**
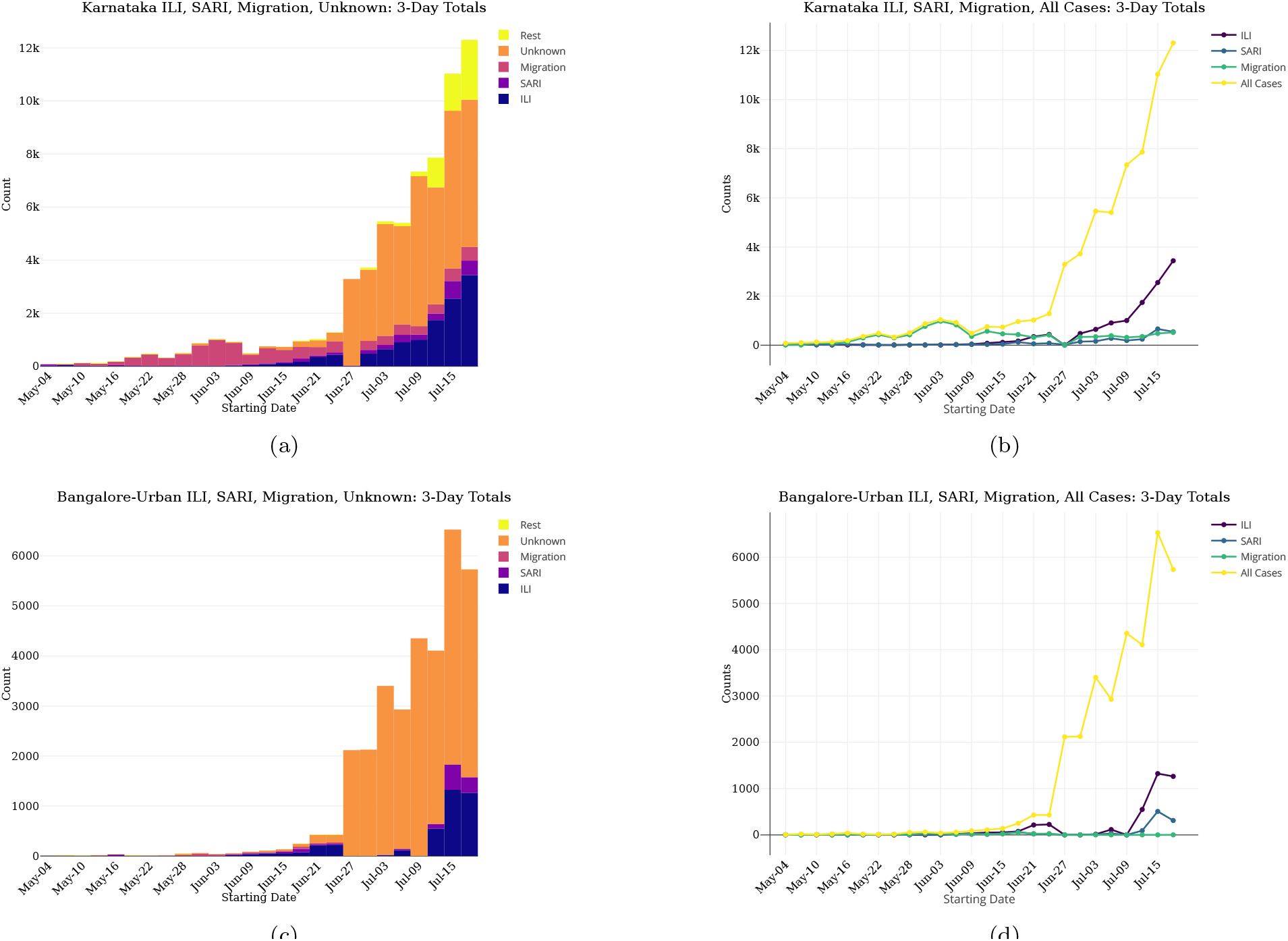
In the graphs on the left, we have considered the sum of daily counts of cases every three days-starting from 4^th^ May and ending on 20^th^ July for Karnataka (Figure 22a and Figure 22b)and Bangalore-Urban (Figure 22c and Figure 22d). The graphs are stacked histograms depicting the proportions of ILI, SARI, Migration, Unknown and the rest of the cases when taken as three day- sums. The bar representing 27^th^, 28^th^ and 29^th^ June is filled with Unknown cases entirely due to the absence of patient description. We see that the proportion of ILI and Unknown cases has been increasing in July. In the graphs on the right, we have again considered the sum of daily counts of cases every three days for Karnataka and Bangalore-Urban. There is dip in these three types of cases on the point representing 27^th^, 28^th^ and 29^th^ June. For Karnataka, it can be seen that from 4^th^ May to 14^th^ June the proportion of Migration cases is quite high, but recently it has decreased. This is clearly not the case for Bangalore-Urban.

**Figure 23:**
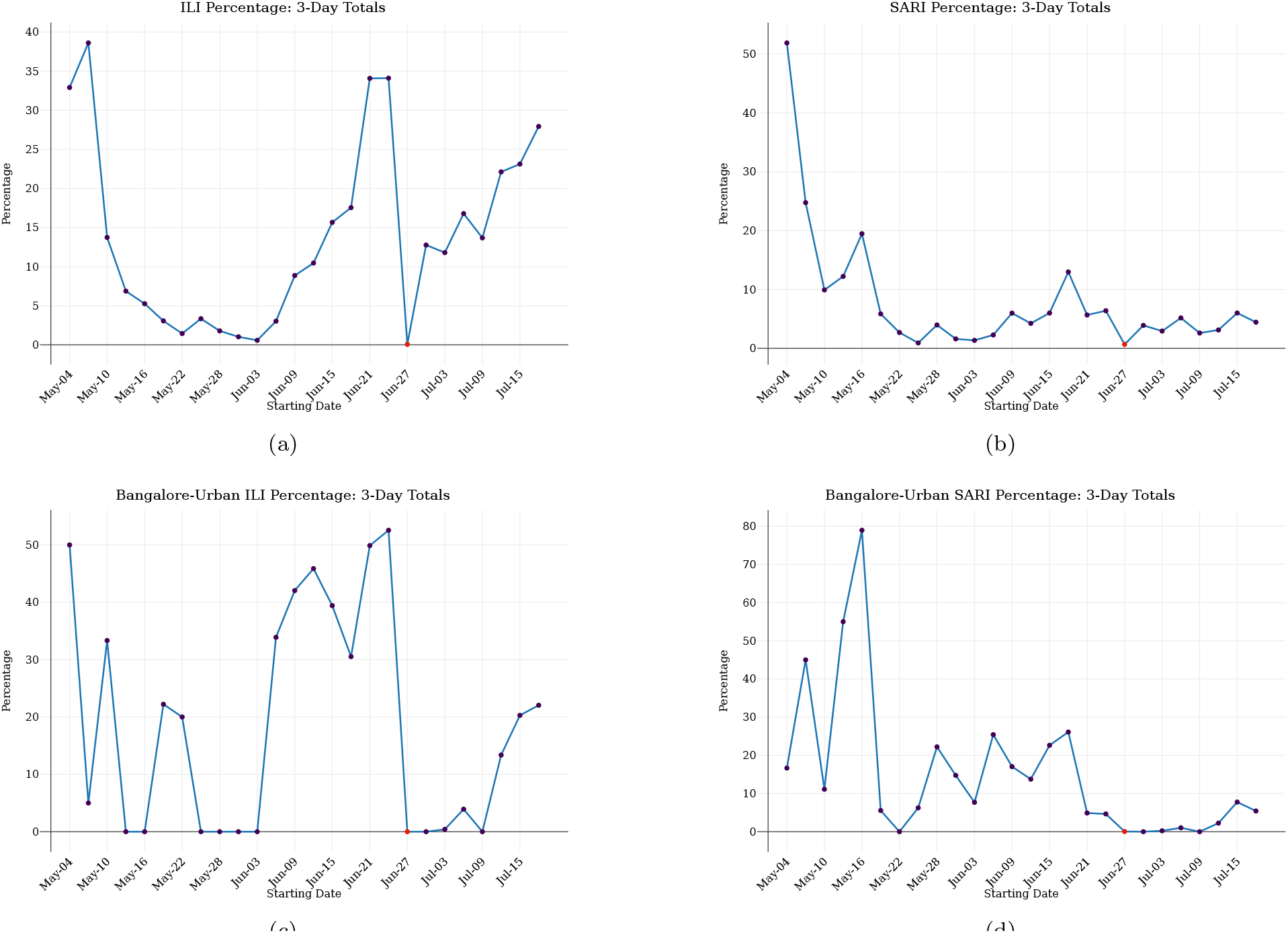
We have considered the sum of daily counts of cases every three days-starting from 4^th^ May and ending on 20^th^ July in Karnataka and Bangalore-Urban. In the above graphs, we have plotted the percentage of cases in “ILI” and “SARI” clusters out of the total cases in Karnataka (Figure 23a and Figure 23b) and Bangalore-Urban respectively (Figure 23c and Figure 23d, taken as the sum of counts for every three days. The red dot denotes the three-day period of 27^th^, 28^th^ and 29^th^ June, when description of patients was absent. This explains the sudden dip in the graphs at the red point.

**Figure 24:**
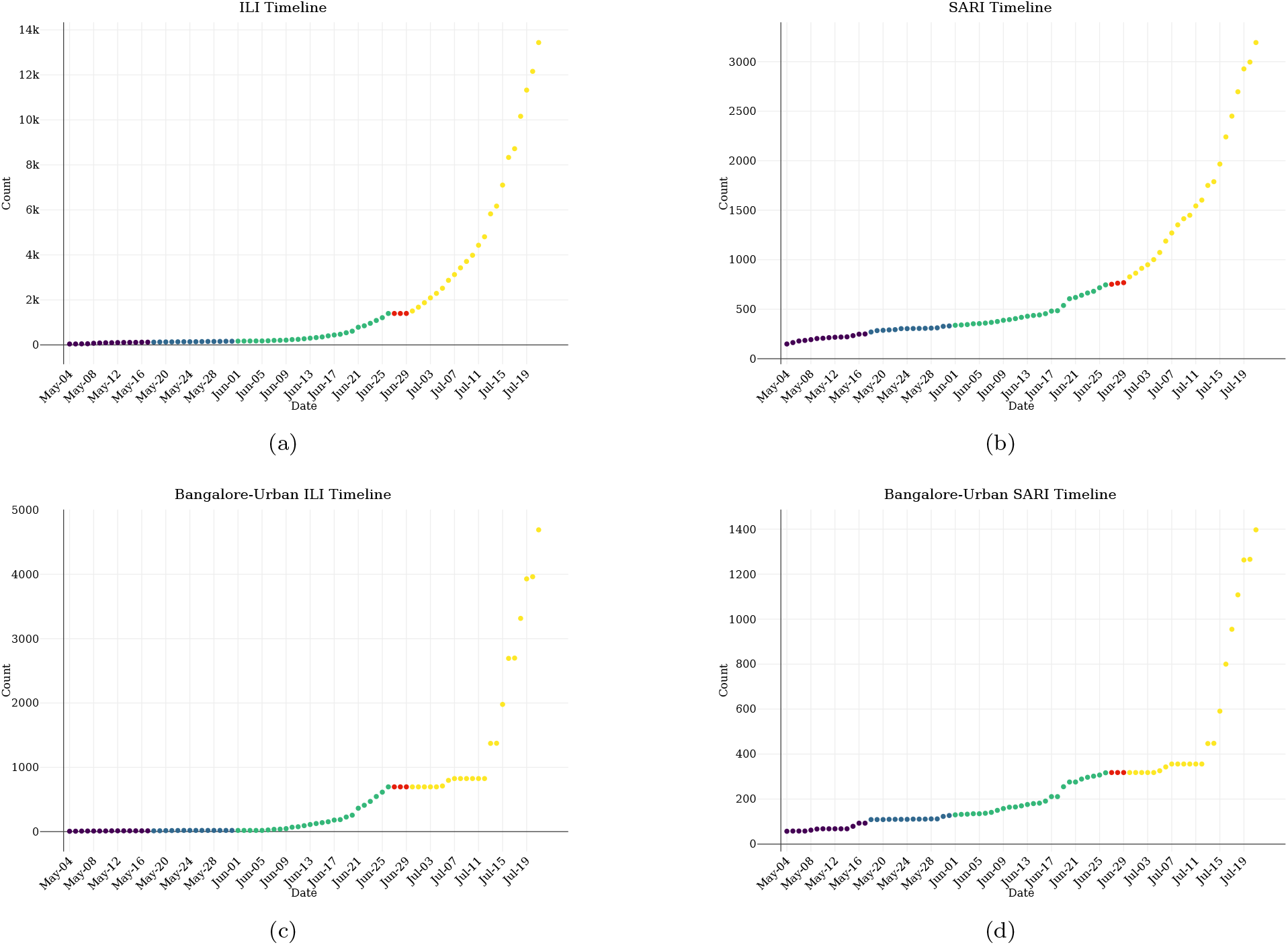
The graphs show the cumulative counts of the “SARI” and “ILI” cluster from 4^th^ May to 21^st^ July in Karnataka (Figure 24a and Figure 24b) and Bangalore-Urban (Figure 24c and Figure 24d). The graphs are color-coded according to the lockdown phase. The 3 red dots visible on the graphs correspond to the dates- 27^th^, 28^th^ and 29^th^ June, when the description of patients was absent. This explains the flat line where the graphs are red. Moreover, till 12^th^ July, the counts seem to increase marginally. Hence the counts afterwards should be considered as a lower bound on the number of ILI and SARI cases and one must note the trend instead of believing the absolute counts where the graph is flat. It can be seen that the counts of ILI reached 14,000 for Karnataka (4700 for Bangalore-Urban) towards the end, while the SARI counts reached 3000 for Karnataka (1400 for Bangalore-Urban).

**Figure 25:**
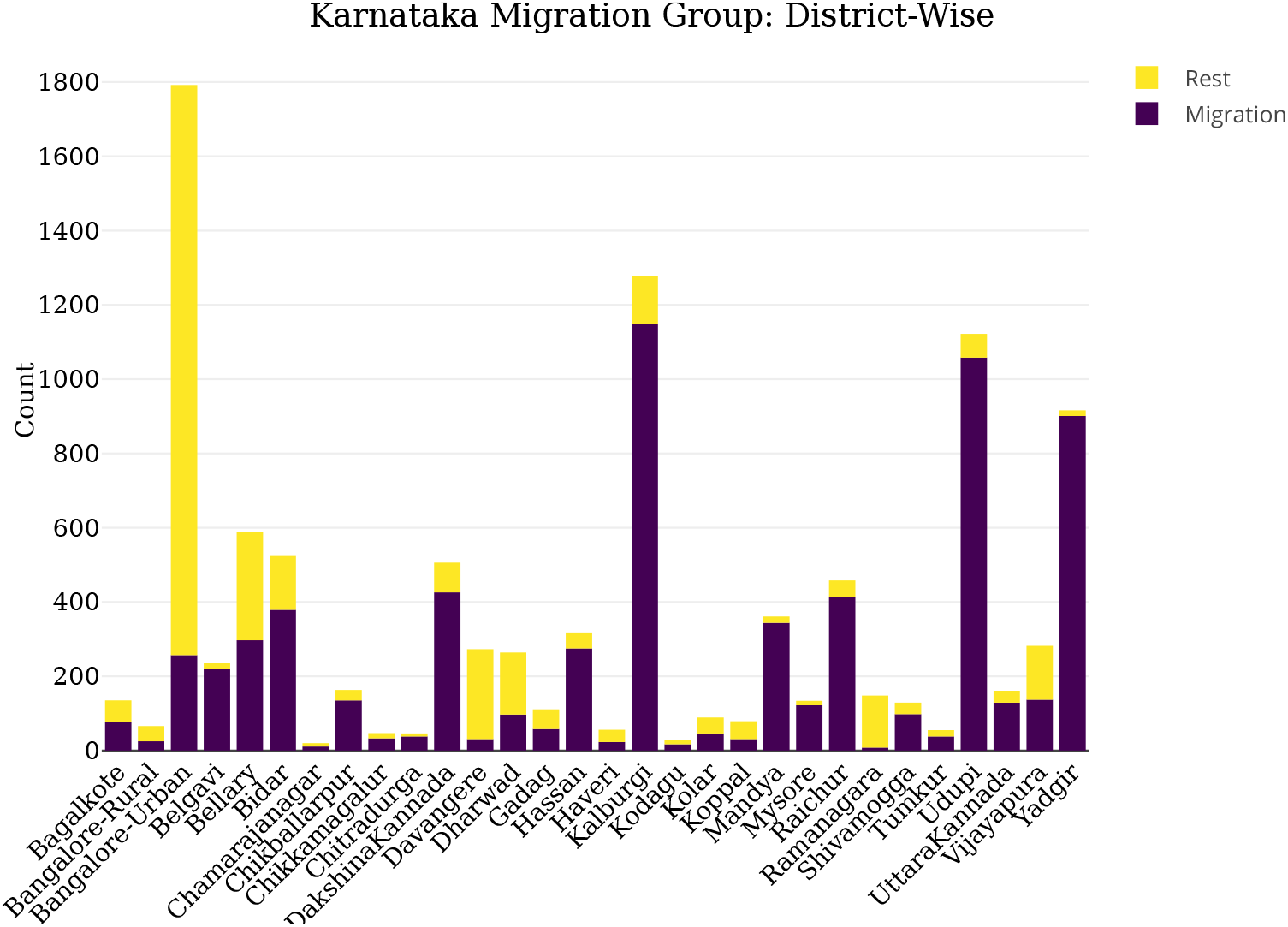
This graph shows district-wise distribution of cases in Karnataka from 4^th^ May to 26^th^ June. The height of bar is the number of cases for the corresponding district. Each bar is divided into number of cases due to migration and cases due to other reasons. Even though the bar for Bangalore-Urban is high, the proportion of migration cases is very less. In contrast, Kalaburagi, Udupi, Yadgir, Mandya, Dakshina Kannada and many others have a high proportion of migration cases

**Figure 26:**
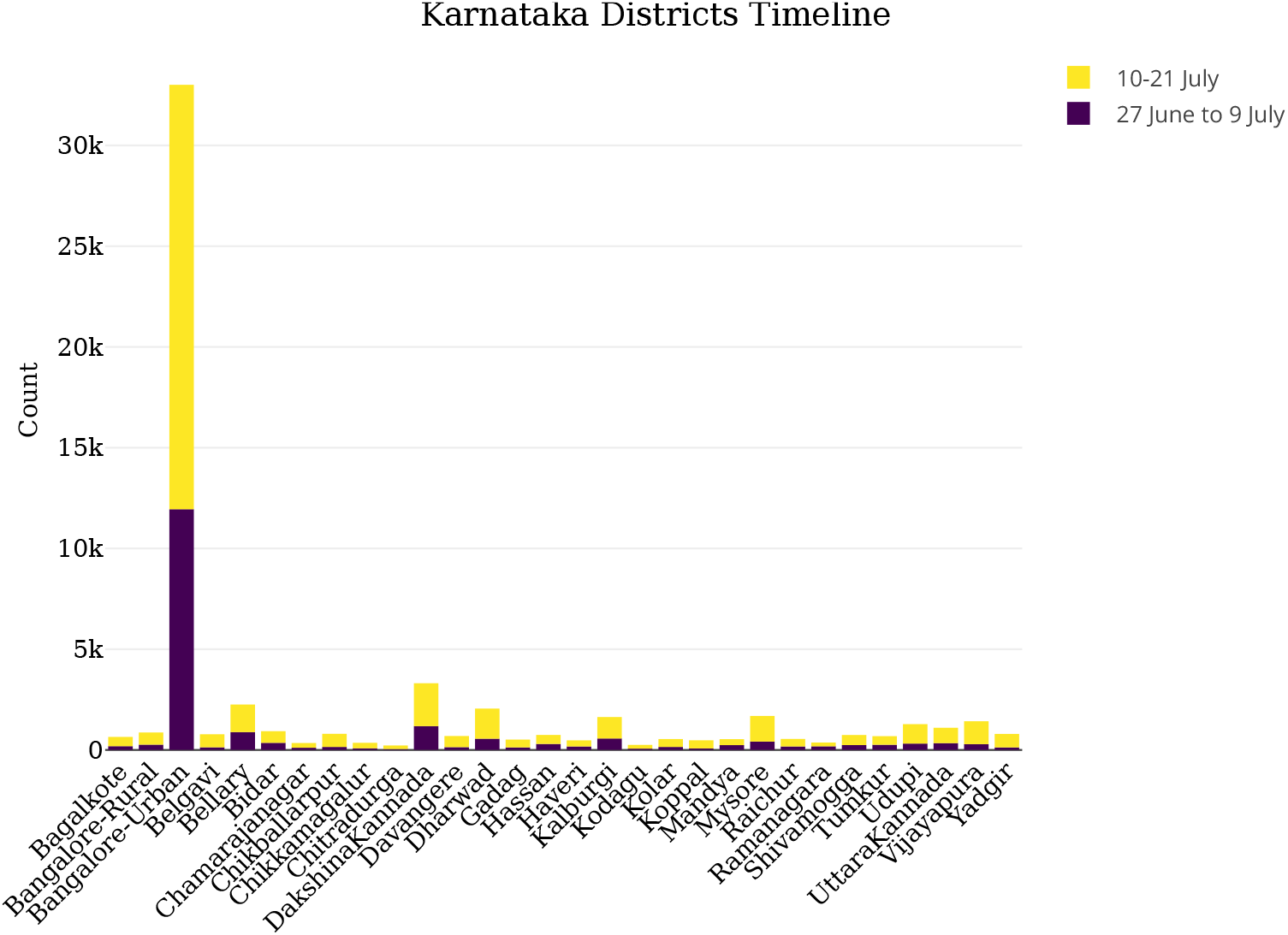
This graph shows district-wise cases in Karnataka from 27th June to 21st July. The height of the bar represents number of cases. We have divided this period into two halves and the bars are color-coded according to each half.

**Figure 27:**
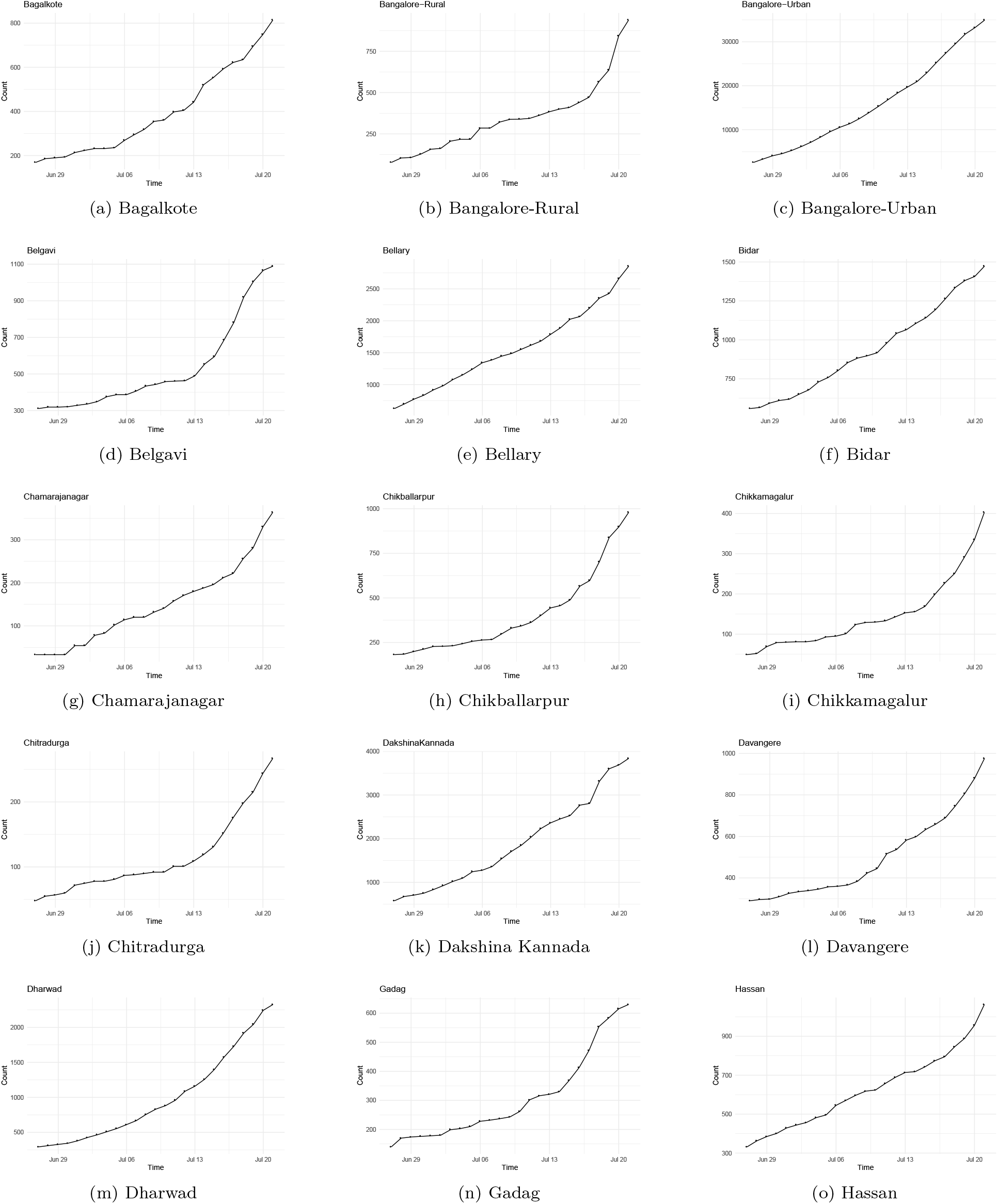
These graphs show the infection timelines of the districts of Karnataka from 27th June to 21st July.

**Figure 28:**
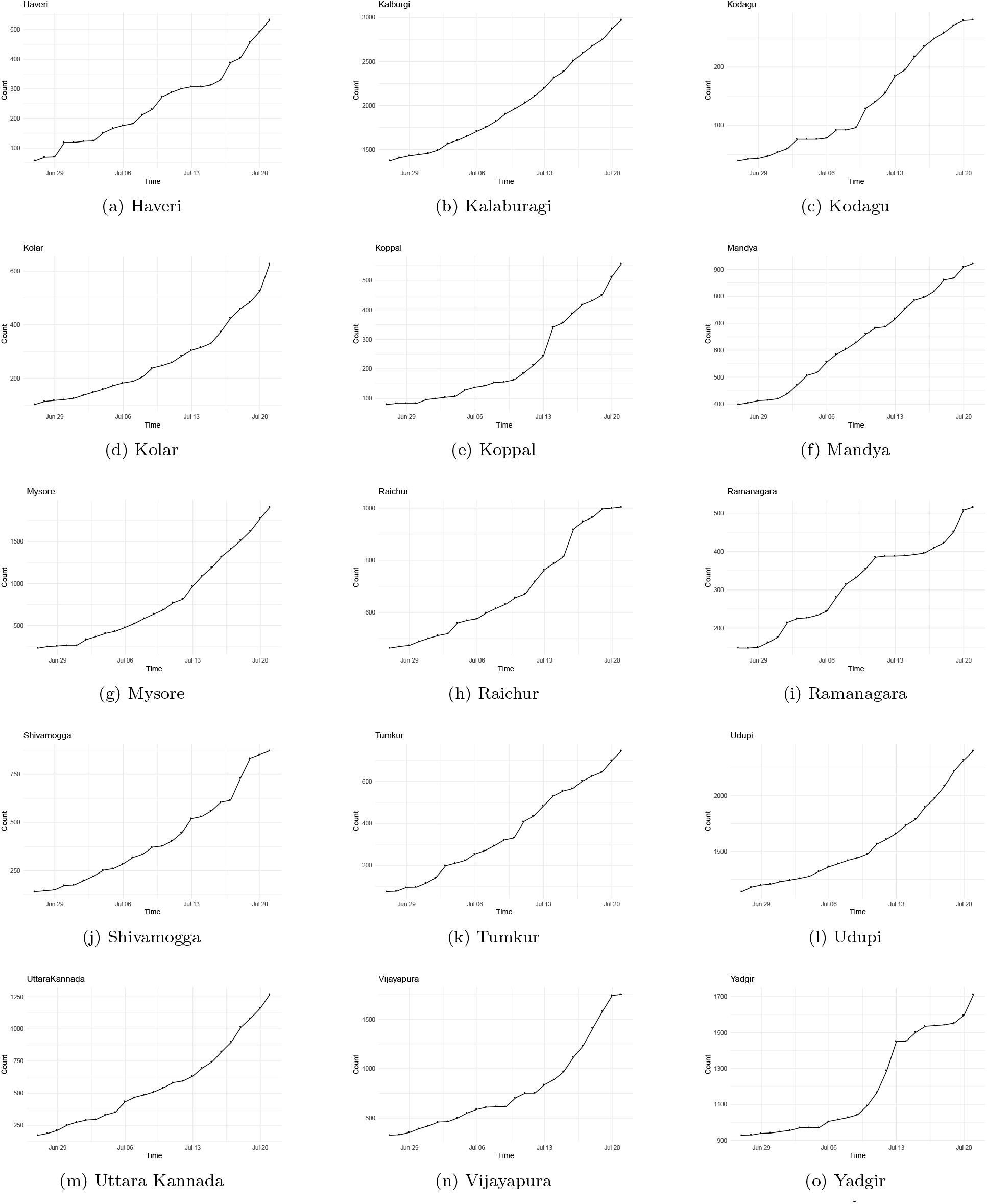
These graphs show the infection timelines of the districts of Karnataka from 27^th^ June to 21^st^ July.

We did notice that in the analysis of “The 8” clusters that “SARI” and “ILI” clusters had *R*^eff^ less than 1 but there was a regular addition of new parents in these clusters. The continuing growth of these clusters indicates presence of viral load in the population. This could be due to one of many reasons. Patients in “SARI”, “ILI and “Unknown” clusters where not entirely contact traced or as their infection source was unknown there were significantly many silent spreaders who did not fall into the contact tracing network. Another aspect to be considered is the Testing policy that was followed in May and June. Ther could have been variations over time such as: non-uniform testing of the population across districts (e.g. testing only on migrants in Phase 3 and Phase 4 due to capacity constraints); and COVID-19 area contact-workers [Health, Law and order, Sanitation] in earlier months were not being tested enough that they inadverdently were spreading the virus. We have already noted that the districts affected most by migration, are Kalaburagi, Udupi and Yadgir (see Figure 25 and above). From Figure 22 we also note the Migration group during the end of June and July did not account for a significant proportion of cases and the current surge in was driven sharply by the cases in Bangalore-urban district. Thus it seems unlikely that the Migration group in Phase 3, Phase 4 and Unlockdown 1.0 is the reason for the surge.

## Data Availability

We have sourced all our data from the Daily Media Bulletins of Government of Karnataka: https://karnataka.gov.in/common-10/en (till April 27th, 2020) and https://covid19.karnataka.gov.in/govt\_bulletin/en (post April 27th, 2020).

https://www.isibang.ac.in/~athreya/incovid19/data.html

## 4 Data

We have sourced all our data from the Daily Media Bulletins of Government of Karnataka: https://karnataka.gov.in/common-10/en (till April 27th, 2020) and https://covid19.karnataka.gov.in/govt bulletin/en (post April 27th, 2020). The media bulletins were very detailed and contained the following information:

- From 9^th^ March to 26^th^ June, the Media Bulletins contained detailed information on the patients. For the 11000 odd cases in this time, information regarding travel history or contact with earlier patients or history of respiratory illness was known.
- The media bulletins stopped containing trace history information for three days 27^th^, 28^th^ and 29^th^ June. For the 3200 odd cases confirmed in this period, only their age, sex and city was known.
- ‹ From 30^th^ June till 11^th^ July, the trace history resumed to it’s former state for all the districts except Bangalore-Urban. For the 12000 odd cases in Bangalore-Urban during this period, the Media Bulletins listed ’Contact Under Tracing’ in the History column for all these patients.
- From 12^th^ July to 21^st^ July, the Media Bulletins contained very consolidated trace history information for all the districts. For the 39500 odd patients during this period, information of the history was either of their Domestic/International travel (but no further details), being a contact (but no Patient ID of the contact was specified), or having ILI or SARI. Information on the ages, sex and city was as earlier.
- 22^th^ July onwards, there was no patient list published in the Media Bulletins.

We have converted them from their pdf format into usable CSV format and made them publicly available for use at our Data Repository at https://www.isibang.ac.in/athreya/incovid19/data.html. We have organised the bulletins into the following three files:

- Karnataka Trace History: This csv file contains information on each patient (age, sex, city, history) who was confirmed before 21st July.
- Karnataka Hospitalization information: This csv file contains information on the patients regarding their hospitalization. The entries in the cells are either H [implying they were hospitalized on that day], C [Cured], D [Deceased], ICU [required an ICU], ICUO [ICU and required Oxygen] or ICUV [ICU and required Ventilator support]. The ICU information is available only till 8^th^ May.
- Karnataka Hospitalization information – Consolidated: This csv file contains consolidated counts of the total counts of Active, Recovered, Discharged and ICU patients.
- Karnataka Testing information: This csv file contains information on the testing and screening done by the Karnataka government.

## Acknowledgements

We would like to thank Gautam Menon for introducing us to the question on Dispersion and also for pointing us to [5]. We would like to thank Rajesh Sundaresan for providing us detailed suggestions that improved the presentation of the paper. Further we would like to thank Biswadeep Karmakar, P. Shankar, Rajesh Sundaresan, Deepayan Sarkar, Mohan Delampady for feedback and discussions.

## Appendix A Model

Let the random variable *ν* represent the number of infections caused by a particular infected individual, called the individual infectiousness. *ν* follows a probability distribution whose mean we will designate as
*R*_eff_. We will assume that

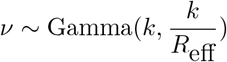

for some *k >* 0 and

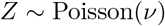

allowing *Z* represents the number of secondary infections caused by each infected individual. A standard calculation shows that for *z* = 0, 1, 2, 3*,…*

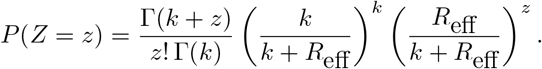

Thus one interprets *Z* as having Negative Binomial distribution with mean *R*_eff_ and Dispersion *k*^1^. It can also be seen that *Z* has variance 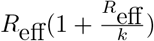. Thus smaller the value *k*, it shall indicate larger variance. Depending on the heterogeneity different models can also be chosen. If one assumed *ν* = *R*_eff_, then we are assuming a homogeneous population where each individual has the same infectiousness. This will imply *Z ∼* Poisson(*R*_eff_) (*k* = *∞*) and if we set (*k* = 1) then *ν ∼* Exponential(*R*_eff_), which arises from mean field models assuming uniform infection and recovery rates, will imply *Z ∼* Geometric(*R*_eff_)

## Appendix B Maximum Likelihood Estimate

Given Data 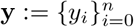 , the log-likelihood (modulo constant terms) is

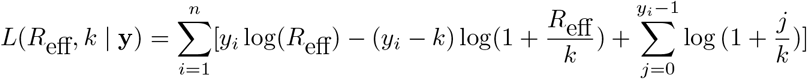

We follow [5] to estimate 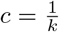. First we rewrite the (conventionally accepted) log-likelihood as a function of *R*_eff_ and 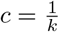.

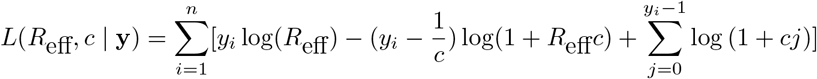

It is then standard (See [9]) that the Maximum Likelihood Estimator for *R*_eff_ is the sample mean, i.e.

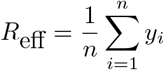

and Maximum Likelihood Estimator for *c* is a solution to

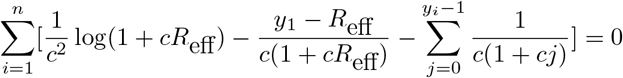

Using (B.1) it is not possible to solve for *c* explicitly. A numerical approximation scheme is used to obtain an approximate value of *c*. We use the uniroot function in R.

## Appendix C *χ*^2^-goodness of fit test

Given Data 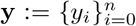. Let 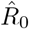 and dispersion 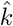 be Maximum likelihood estimators. To see if Negative Binomial with mean 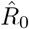 and dispersion 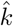 is a good fit for the data y we shall perform the χ^2^-goodness of fit
test. We will consider the range to

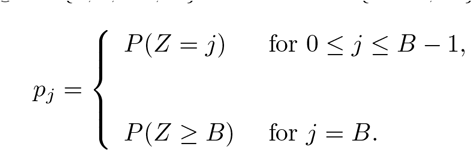

Let y_1_, y_2_…,yn be the offspring data from a given cluster and let

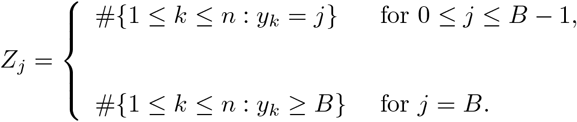

Then consider the statistic

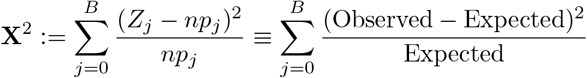

As we have estimated two parameters, it is known that X^2^- has *χ*^2^_*B−*2_ degrees of freedom, asymptotically as *n → ∞*. One way to test if *Z* is the correct fit for the cluster is to compute the

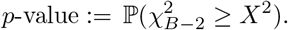

There is strong evidence against the possibility that data arose from that model if *p*-value is very small.

## Appendix D Confidence Intervals

To compute the confidence interval for the negative binomial dispersion parameter *k*, we compute it for its reciprocal *c* and then invert it. We noted earlier that the maximum likelihood estimate for *c* had to be solved numerically and it is known that the asymptotic sampling variance is given by a series expansion (See [9]). Let 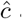 and 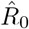 be the M.L.E. obtained. Then let

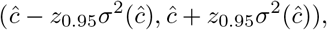

Then the variance of 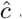 is given by

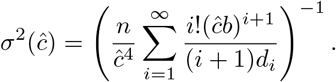

The 95% confidence interval for c is then given by

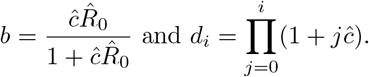

with *z*_0.95_ being the 95^th^ percentile of the standard normal distribution. The 95% confidence interval for *k* is then given by

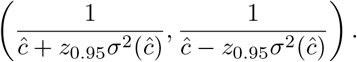

Note that the above interval will not be symmetric around *k* due to the inversion. For the computation of Variance in (D.1) we use a tolerance of 10^*−*10^.

## Footnotes

1 In the Epidemiological literature *k* is referred to as Dispersion and *k >* 0 is assumed, while in the Statistics literature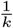 is referred to as Dispersion given the connection with the Gamma distribution and is allowed to take negative values up to 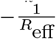

